# Subgenomic and negative sense RNAs are not markers of active replication of SARS-CoV-2 in nasopharyngeal swabs

**DOI:** 10.1101/2021.06.29.21259511

**Authors:** Anthony Chamings, Tarka Raj Bhatta, Soren Alexandersen

## Abstract

Severe acute respiratory syndrome coronavirus 2 (SARS-CoV-2) has spread rapidly in the global population since its emergence in humans in late 2019. Replication of SARS-CoV-2 is characterised by transcription and replication of genomic length RNA and shorter subgenomic RNAs to produce virus proteins and ultimately progeny virions. Here we explore the pattern of both genome-length and subgenomic RNAs and positive and negative strand SARS-CoV-2 RNAs in diagnostic nasopharyngeal swabs using sensitive probe based PCR assays as well as Ampliseq panels designed to target subgenomic RNAs. We successfully developed a multiplex PCR assay to simultaneously measure the relative amount of SARS-CoV-2 full length genomic RNA as well as subgenomic N gene and subgenomic ORF7a RNA. We found that subgenomic RNAs and both positive and negative strand RNA can be readily detected in swab samples taken up to 19 and 17 days post symptom onset respectively, and are strongly correlated with the amount of genomic length RNA present within a sample. Their detection and measurement is therefore unlikely to provide anymore insight into the stage of infection and potential infectivity of an individual beyond what can already be inferred from the total viral RNA load measured by routine diagnostic SARS-CoV-2 PCRs. Using both an original commercial and two custom SARS-CoV-2 Ampliseq mini-panels, we identified that both ORF7a and N gene subgenomic RNAs were consistently the most abundant subgenomic RNAs. We were also able to identify several non-canonical subgenomic RNAs, including one which could potentially be used to translate the ORF7b protein and others which could be used to translate ORF9b and the ORF N* which has arisen from a new transcription regulatory sequence recently created by mutations after SARS-CoV-2 jumped into people. SARS-CoV-2 genomic length and subgenomic length RNA’s were present in samples even if cellular RNA was degraded, further indicating that these molecules are likely protected from degradation by the membrane structures seen in SARS-CoV-2 infected cells.

## Introduction

Severe acute respiratory syndrome coronavirus 2 (SARS-CoV-2), the causative agent of human coronavirus disease 2019 (COVID-19), is a novel betacoronavirus which was first detected in humans in late 2019^[1,2]^. The virus was readily transmittable from person to person and rapidly spread worldwide causing unprecedented economic and social disruption in many countries^[3]^. While infection can result in severe life-threatening respiratory disease and death in some individuals, a large proportion of people will be asymptomatic or only exhibit mild respiratory signs^[1,4]^. Therefore, knowing who is infected and when they may possibly be infectious cannot be done based on clinical symptoms alone^[5]^. The most commonly used method to determine the infection status of an individual is the real-time reverse transcription PCR (rRT-PCR)^[6]^. The rRT-PCR detects and measures the amount of SARS-CoV-2 RNA within a sample. Some individuals can test positive for virus RNA for 3 weeks or more^[6]^ after they were thought to be initially infected, and long after the time when infectious virus shedding is thought to cease which is around 7-8 days post detection of symptoms in most people^[7-9]^. Therefore detection of the presence of viral RNA alone may not necessarily indicate that an individual is infectious. The gold standard method for detecting whether infectious virus is being excreted is inoculation of susceptible cell cultures^[10]^. However this is not practical in most diagnostic laboratories, and importantly, may not be sensitive enough to detect low levels of potential infectivity^[7,11]^. This has led to a number of other molecular indicators such as detection of subgenomic RNAs or negative sense RNA being proposed as possible markers of whether an individual is likely to be infectious or not to aid in the control of SARS-CoV-2^[7,12-14]^. The rationale being that these molecules are generated intracellularly by the virus during virus replication. SARS-CoV-2, like other coronaviruses, is an enveloped virus with a single stranded positive sense RNA genome of nearly 30,000 nucleotides. The genomic RNA consists of a 5’ UTR, two large open reading frames (ORFs) - ORF1a and ORF1b, which occupy two thirds of the 5’ end of the genome and which express two large polyproteins which are proteolytically cleaved into 16 known non-structural proteins, and several ORFs which encode the structural and accessory proteins needed for the virus to replicate/transcribe and produce progeny virions, and a polyadenylated 3’ UTR^[15]^. After initial infection, coronavirus replication inside a cell is believed to involve initial translation of the ORF1ab proteins from the genomic RNA and the formation of subcellular organelles including convoluted membranes (CM) and double membrane vesicles (DMV)^[16-19]^. Production of negative stranded RNA, double stranded RNA intermediates and new positive stranded genomic and subgenomic coronavirus RNAs occurs within replication transcription complexes associated with these membrane structures as part of the process of coronavirus RNA replication and transcription^[19]^. These membrane structures are currently understood to concentrate virus proteins and RNA, provide the framework on which RNA synthesis can take place and possibly shield the virus replication complexes and RNA from cellular defences ^[16,20]^. These structures have also been observed in SARS-CoV-2 infected cells by electron microscopy^[21]^ and therefore the mechanisms of SARS-CoV-2 replication are likely similar to other coronaviruses.

Within these membrane structures, negative strand RNA, which most likey exists as partial or complete double stranded RNA^[16,19]^, serves as a template for new positive strand copies of the genomic length RNA (gRNA) to be used as either mRNA for additional production of virus proteins or to be packaged inside progeny virus particles. A number of shorter subgenomic RNAs are also produced via a complex method of discontinuous negative strand RNA synthesis serving as templates for generation of positive strand RNA copies which serve as messenger RNAs used to express each of the structural and accessory proteins of SARS-CoV-2^[15,22]^. Each subgenomic RNA molecule shares a common leader sequence of approximately 65-69 nucleotides within the 5’UTR of the SARS-CoV-2 genome^[23]^. During negative strand RNA synthesis, the viral RNA polymerase pauses at a transcription regulatory sequences (TRS) upstream from the ORFs responsible for encoding the structural and accessory proteins within the 3’-third of the genome. The nascent RNA is then joined to the TRS within the leader sequence creating a negative sense subgenomic RNA which is used as a template to make positive sense subgenomic RNAs as mentioned above^[22]^.

Several studies have proposed using subgenomic RNA or negative sense SARS-CoV-2 RNA as a marker of active replication of the virus within an individual^[7,11,13,14,24]^. However, we recently reported that we were able to detect subgenomic RNA and negative sense RNA in samples up to 17 days post infection, and that subgenomic RNAs are relatively stable and likely persist in samples protected by the double membrane structures created during the replication of the virus in the cytoplasm of SARS-CoV-2 infected cells^[23]^. This theory is supported by studies in other coronaviruses such as mouse hepatitis virus, where double membrane vesicles containing double stranded RNA have been observed in cells late in the infection cycle and which are not associated with active replication^[20]^. Other authors have similarly suggested that the amount of subgenomic RNA is simply related to the amount of total SARS-CoV-2 RNA present within a sample, and correlates poorly with the shedding of infectious virus^[9,25]^. Therefore subgenomic RNAs, or the presence of any negative strand RNA, may not necessarily indicate that virus replication is currently occurring, only that replication of SARS-CoV-2 has occurred at some point in the recent past^[23]^.

Given that there is ongoing interest in subgenomic RNAs as a potential marker of active replication and a dichotomy of conclusions being drawn within the scientific community as to the utility of subgenomic RNA as a marker of infectivity^[7,9,11,13,14,23,24]^, we decided to further examine the presence of subgenomic RNAs in a larger number of routine diagnostic SARS-CoV-2 positive nasopharyngeal swab samples. These swabs were collected subsequent to our last study during the second SARS-CoV-2 epidemic wave in Victoria, Australia in mid-to late 2020^[23,26]^. In addition to including more routine diagnostic samples, we also developed and used more sensitive molecular assays to measure relative loads of subgenomic RNAs as well as negative and positive strand SARS-CoV-2 RNA. We used these tools to investigated if there was any relationship between subgenomic RNA, genomic RNA, positive and negative strand RNA, sample characteristics including the time of onset of clinical symptoms and the quality of cellular RNA.

## Results

### Development of a multiplex RT-PCR assay to detect SARS-CoV-2 full length genomic RNA, subgenomic ORF7a RNA and subgenomic N Gene RNA

We had previously identified by amplicon-based sequencing that subgenomic N and ORF7a RNA were frequently the most abundant subgenomic RNAs present in naso-oropharyngeal swabs from infected individuals^[23]^. Based on those findings, we designed probe based PCR assays to quantitate the levels of these two subgenomic RNAs along with a 5’UTR PCR to quantify the amount of full-length genomic RNA in samples. We also developed a total ORF7a probe based PCR to quantitate the total amount of SARS-CoV-2 genomic RNA and subgenomic RNAs for S, ORF3a, E, M, ORF6 and ORF7a RNA molecules present in a sample.

We selected 24 positive SARS-CoV-2 nasopharyngeal swabs from 16 individuals collected during 2020 from two diagnostic laboratories in Victoria Australia (Table 1). We first tested the PCRs as single target assays, and demonstrated that the assays could successfully detect SARS-CoV-2 genomic length RNA with both the 5’UTR and ORF7a total PCRs in 23 out of 24 positive swabs. Subgenomic ORF7a RNA and subgenomic N gene RNAs were detected in 17 and 22 out of 24 swabs with their respective assays (Supplementary Table S1 and Supplementary Fig. S1). We then attempted duplexing the 5’UTR and subgenomic ORF7a PCRs (Supplementary Table S2 and Supplementary Fig. S2). Again 23 out of 24 swabs were positive on the 5’UTR assay and 16 out of 24 were positive for the subgenomic ORF7a RNA. The SARS-CoV-2 infected cell culture was positive for the single target and duplex assays and all SARS-CoV-2 negative swabs were negative.

**Table 1.** Table showing summary information about the individuals and samples included in this study. Samples from a total of 17 individuals, including one SARS-CoV-2 negative control individuals (Individual 1). A pool of 10 SARS-CoV-2 negative swabs was also used as a negative PCR control. Swab sample identification (ID), clinical symptoms onset and description of clinical symptoms where available, sampling date and the results of the initial diagnostic SARS-COV-2 RT-PCR test (Ct value) are provided.

| Individual ID | Gender | Age group (Years) | Swab Sample ID | Epidemic wave sample taken† | Days since onset of symptoms/Initial swab | Clinical Symptoms at swab collection | Hospitalized | Initial SARS-CoV2 RT-PCR result (Ct Value) | Comments |
| --- | --- | --- | --- | --- | --- | --- | --- | --- | --- |
| 1 | F | 20-40 | GC-28 | 1 | 0* | Fever, cough sore throat | N | Not detected | Negative control swab sample used in PCR assays |
| 2 | F | 20-40 | GC-26 | 1 | 7 | Sore throat, dry cough | N | Detected (21) |  |
| 3 | F | 20-40 | GC-13 | 1 | 4 | Body aches, headache, dry cough, shortness of breath | N | Detected (29) |  |
| 4 | F | 40-60 | GC-24 | 1 | 14* | Cold, sinusitis |  | Detected (31) |  |
| 6 | F | 40-60 | GC-14 | 1 | 0 | Unspecified | N | Detected (18) |  |
| 6 |  |  | GC-23 |  | 11 | Asymptomatic |  | Detected (31) |  |
| 6 |  |  | GC-51 |  | 17 | Asymptomatic |  | Detected (31) |  |
| 7 | F | 40-60 | GC-21 | 1 | 16 | Sore throat, shortness of breath, runny nose | Y | Detected (31) |  |
| 7 |  |  | GC-58 |  | 47 | Not specified |  | Detected (38) |  |
| 8 | M | 40-60 | GC-25 | 1 | 2 | Sore throat, hoarse voice | N | Detected (19) |  |
| 11 | F | 80- | GC-242 | 2 | 0 | Not specified | N | Detected (28) |  |
| 11 |  |  | GC-290 |  | 11 | Not specified |  | Detected (34) |  |
| 12 | F | 60-80 | GC-251 | 2 | 0 | Not specified | N | Detected (17) |  |
| 12 |  |  | GC-288 |  | 10 |  |  | Detected (34) |  |
| 13 | F | 40-60 | GC-277 | 2 | 0 | Asymptomatic | N | Detected (21) |  |
| 13 |  |  | GC-329 |  | 19 |  |  | Detected (34) |  |
| 14 | F | 80- | GC-238 | 2 | 0 | Not specified |  | Detected (25) |  |
| 14 |  |  | GC-291 |  | 13 |  |  | Detected (22) |  |
| 16 | F | 80- | GC-366 | 2 | 14 | Not specified | Y | Detected (30) |  |
| 16 |  |  | GC-365 |  | 17 |  |  | Detected (33) |  |
| 17 | F | 40-60 | GC-316 | 2 | 1 | Fever, runny nose, headache | N | Detected (20) |  |
| 18 | M | 60-80 | GC-310 | 2 | 17 | Cough, shortness of breath | Y | Detected (23) |  |
| 19 | M | 40-60 | GC-199 | 2 | 1 | Fever, cough, shortness of breath,<br>headache | Y | Detected (24) |  |
| 20 | F | 80- | GC-292 | 2 | 10 | Not specified | Y | Detected (18) |  |
| 21 | F | 40-60 | GC-243 | 2 | 0 | Not specified | N | Detected (24) |  |
| Negative<br>Pool 1 |  |  |  | 1 |  | Pool of 10 individuals |  | Not detected | This pool was used as a<br>negative control for the<br>PCR assays |

\*Actual onset of symptoms was not recorded for these samples, so we counted the days from the time of initial swab collection. † 1 - first epidemic wave in Victoria Australia during March-May 2020 and 2 - second epidemic wave in Victoria Australia July-September 2020. Negative pools 1 was made from SARS-CoV2 negative individuals tested during the first wave of SARS-CoV-2 infections.

We then triplexed the subgenomic N PCR with the 5’UTR and subgenomic ORF7a assays. We were able to detect the SARS-CoV-2 5’UTR in 23 out of 24 known SARS-CoV-2 positive swab samples, the subgenomic ORF7a in 19 and the subgenomic N gene is 21 out 24 SARS-CoV-2 positive swabs (Table 2 and Fig. 1). The cell culture sample was positive for each target in the triplex assay, and the negative swabs remained negative. The 5’UTR assay in triplex was on average 1.6 Ct’s (IQR: -1 to - 2.1 Ct’s) lower than the single target 5’ UTR and now very closely approximated the values seen in the single target ORF7a total RNA PCR (average delta Ct: 0.1 Ct’s; IQR: -0.4-0.7 Ct’s). The triplex subgenomic ORF7a assay was on average 1.7 Ct’s lower (IQR: -1.5 to -2.2 Ct’s) than the single target assay. The triplex subgenomic N gene assay was about equal in sensitivity or slightly more sensitive than the single target subgenomic N gene assay (average 0.25 Ct’s lower, IQR: -0.2 to -0.45 Ct’s). The triplex assay was repeatable with an average difference of 0.4 (range: 0 to 1.9), 0.2 (range: 0 to 1.6) and 0.2 Ct’s (range: 0 to 0.8) observed for the replicates of each of the 5’UTR, subgenomic ORF7a and subgenomic N gene PCRs targets respectively. Thus, there was no loss of sensitivity in the multiplex assays relative to the single target assays despite the fact that each multiplex assay shared the same forward primer. In fact, there was a decrease of 1-2 Ct’s of both the 5’UTR and subgenomic 7a assays when multiplexed, and a very slight decrease in the Ct (0.25 Ct’s) of the subgenomic N gene in the triplex assay.

**Table 2.** Ct values obtained from the triplex SARS-CoV-2 genomic and subgenomic PCR assays for the swab samples from the 16 SARS-CoV-2 positive individuals, negative individual and pool and from 48hr virus cell culture supernatant.

| Individual | Swab Sample ID | Days since onset of symptoms/Initial swab | 5'UTR Genomic PCR Repeat 1 | 5'UTR Genomic PCR Repeat 2 | ORF7a subgenomic PCR Repeat 1 | ORF7a subgenomic PCR Repeat 2 | N gene subgenomic PCR Repeat 1 | N gene subgenomic PCR Repeat 2 |
| --- | --- | --- | --- | --- | --- | --- | --- | --- |
| 1 | GC-28 | 0* | NEG | NEG | NEG | NEG | NEG | NEG |
| 2 | GC-26 | 7 | 24.7 | 24.5 | 27.8 | 27.7 | 27.8 | 27.9 |
| 3 | GC-13 | 4 | 27.7 | 28.2 | 31.5 | 31.4 | 30.3 | 30.8 |
| 4 | GC-24 | 14* | 34.2 | 34.2 | NEG | NEG | NEG | NEG |
| 6 | GC-14 | 0 | 17.8 | 18 | 21.3 | 21.5 | 20.4 | 20.5 |
| 6 | GC-23 | 11 | 32.2 | 31.5 | 36.4 | NEG | 33.9 | 34.4 |
| 6 | GC-51 | 17 | 36.3 | 36.9 | NEG | NEG | NEG | NEG |
| 7 | GC-21 | 16 | 35.3 | 33.9 | NEG | NEG | 36.3 | NEG |
| 7 | GC-58 | 47 | NEG | NEG | NEG | NEG | NEG | NEG |
| 8 | GC-25 | 2 | 19.2 | 19.5 | 22.4 | 22.5 | 21.3 | 21.3 |
| 11 | GC-242 | 0 | 26.3 | 26.2 | 28.9 | 28.7 | 27.1 | 27.3 |
| 11 | GC-290 | 11 | 31.8 | 32.2 | 35 | 35 | 34.2 | 33.4 |
| 12 | GC-251 | 0 | 16.5 | 16.5 | 19.3 | 19.4 | 17.9 | 18 |
| 12 | GC-288 | 10 | 33.5 | 34.2 | 34.9 | 35.3 | 35.3 | 35.2 |
| 13 | GC-277 | 0 | 18.7 | 18.8 | 22.6 | 22.5 | 21.6 | 21.6 |
| 13 | GC-329 | 19 | 35.5 | 35 | 37.4 | 35.8 | NEG | 34.8 |
| 14 | GC-238 | 0 | 22 | 21.9 | 27.8 | 27.6 | 26.9 | 26.8 |
| 14 | GC-291 | 13 | 20.4 | 20.7 | 23.6 | 23.8 | 23 | 23.2 |
| 16 | GC-366 | 14 | 34.8 | 34.8 | NEG | NEG | 35.9 | 35.9 |
| 16 | GC-365 | 17 | 34.5 | 36.4 | NEG | 36.1 | 35.4 | NEG |
| 17 | GC-316 | 1 | 17.5 | 17.8 | 20.7 | 20.9 | 19.9 | 19.8 |
| 18 | GC-310 | 17 | 21.3 | 21.3 | 25 | 25 | 24.2 | 24.1 |
| 19 | GC-199 | 1 | 22.5 | 22.6 | 26.5 | 26.4 | 25.1 | 25 |
| 20 | GC-292 | 10 | 15.9 | 16 | 19.3 | 19.4 | 18.2 | 18.3 |
| 21 | GC-243 | 0 | 22.4 | 22.8 | 25.2 | 25.5 | 23.8 | 24.2 |
| SARS-CoV-2 Negative swab pool | Pool of 10 Neg. swabs | Not specified | NEG | NEG | NEG | NEG | NEG | NEG |
| Cell culture 48Hr |  | 48 hr post inoculation | 16.9 | 17.1 | 20.8 | 21 | 19.6 | 19.8 |

ND: Test not performed. NEG: SARS-CoV-2 not detected.
\*Date of onset of clinical signs not provided. Number of days since the collection of the first nasopharyngeal swabs is shown.

**Figure 1.**
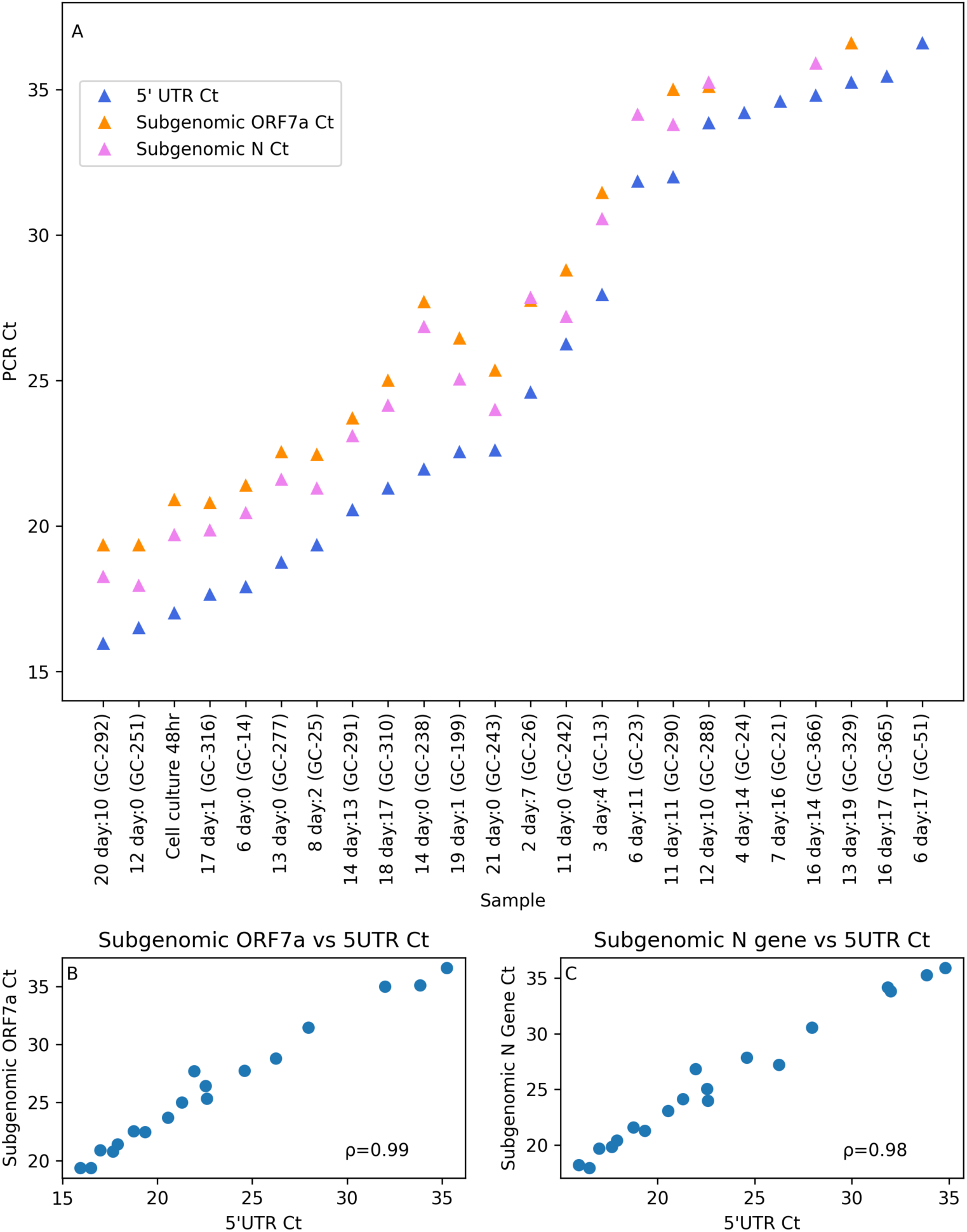
A: The average Ct’s of the replicates of the 5’UTR genomic and ORF7a and N gene subgenomic triplex PCRs for the SARS-CoV-2 positive naso-oropharyngeal swabs and the SARS-CoV-2 48 hr cell culture arranged by ascending 5’UTR Ct (PCR negative samples not shown). B: Correlation plot of the subgenomic ORF7a PCR Ct vs the 5’UTR Ct. C: Correlation plot of the subgenomic N gene PCR Ct vs the 5’UTR Ct. The Spearman ranked correlation coefficient (ρ) is shown in each figure B and C (n=24 data points from 24 samples run twice)

The triplex assay detected SARS-CoV-2 RNA in all but one known positive nasal swab sample. The single false negative sample (GC-58) was a very borderline positive sample with a Ct of 38 when tested by the original diagnostic laboratory, possibly accounting for why it was now testing negative several months after being originally collected. The efficiencies of the three assays when triplexed together were calculated at 95% by testing serial dilutions of PCR amplicons.

### Quantitation of SARS-CoV-2 genomic and 7a and N subgenomic RNAs using probe based real-time PCR assays reveals that the amount of subgenomic RNA is highly correlated to the amount of SARS-CoV-2 full length genomic RNA

Using the Ct’s obtained from the triplex PCR, we quantified the relative amounts of the SARS-CoV-2 full length RNA, as indicated by the 5’UTR RNA target mentioned above, and the ORF7a and N gene subgenomic RNAs in the cell culture and nasopharyngeal samples (Figure 1). It was immediately evident that the Ct’s of the subgenomic assays appeared to follow the Ct of the 5’UTR which measured the amount of full-length genomic RNA in the samples. The subgenomic ORF7a assay was on average 3.1 Ct’s higher than the 5’UTR (IQR: 2.8-3.7 Ct’s) and the subgenomic N gene assay was on average 2.3 Ct’s higher than the 5’UTR (IQR: 1.6-2.7 Ct’s) (Figure 1A). We then plotted the Ct of each subgenomic assay against the Ct of the 5’UTR (Figure 1B and C). The Ct’s of both subgenomic assays were highly correlated with the Ct of the 5’UTR PCR, and the subgenomic ORF7a and N gene reported a Spearman’s rank correlation coefficient (ρ) of 0.99 and 0.98 respectively. Samples with a lot of genomic length RNA (low 5’UTR Ct) had more subgenomic RNA, and samples with a high Ct had less. At a Ct of 30 or higher for the 5’UTR PCR, the detection of one or both of the subgenomic RNAs became inconsistent, presumably as the levels of RNA reached the detection limit of each of the subgenomic PCRs. In all the samples with a 5’UTR PCR Ct of 30 or higher, the subgenomic PCRS, were reporting Cts in the mid-30’s or higher. At these very high Ct’s, it was likely that amplification of the subgenomic targets was beginning to behave stochastically, and therefore accurate measurement of the subgenomic targets in these samples would likely require several repeated PCRs to ensure amplification occurred in at least one of the repeats.

We calculated the ratios of full length genomic RNA to each of the subgenomic RNA molecules by 1.9^(*subgenomic PCR Ct*−5*′UTR PCR Ct*)^(with 1.9 indicating 95% PCR efficiency per cycle). There was thus on average 9.62 more genomic full length SARS-CoV-2 RNA than subgenomic ORF7a RNA (IQR: 6.4 – 10.4) and an average of 4.3 times more full length genomic RNA relative to subgenomic N gene RNA (IQR: 2.9-5.5). With the exception of GC-238 (Individual 14, on the day of symptom onset) and samples with a very high 5’UTR Ct (GC-288, GC-329 and GC-366) where the subgenomic PCRs were reaching their limits of detection, the ratios of all other samples lay within a 2-fold range around the mean value irrespective of whether the sample was collected at the onset of symptoms or more than two weeks later (Figure 2A and B).

**Figure 2.**
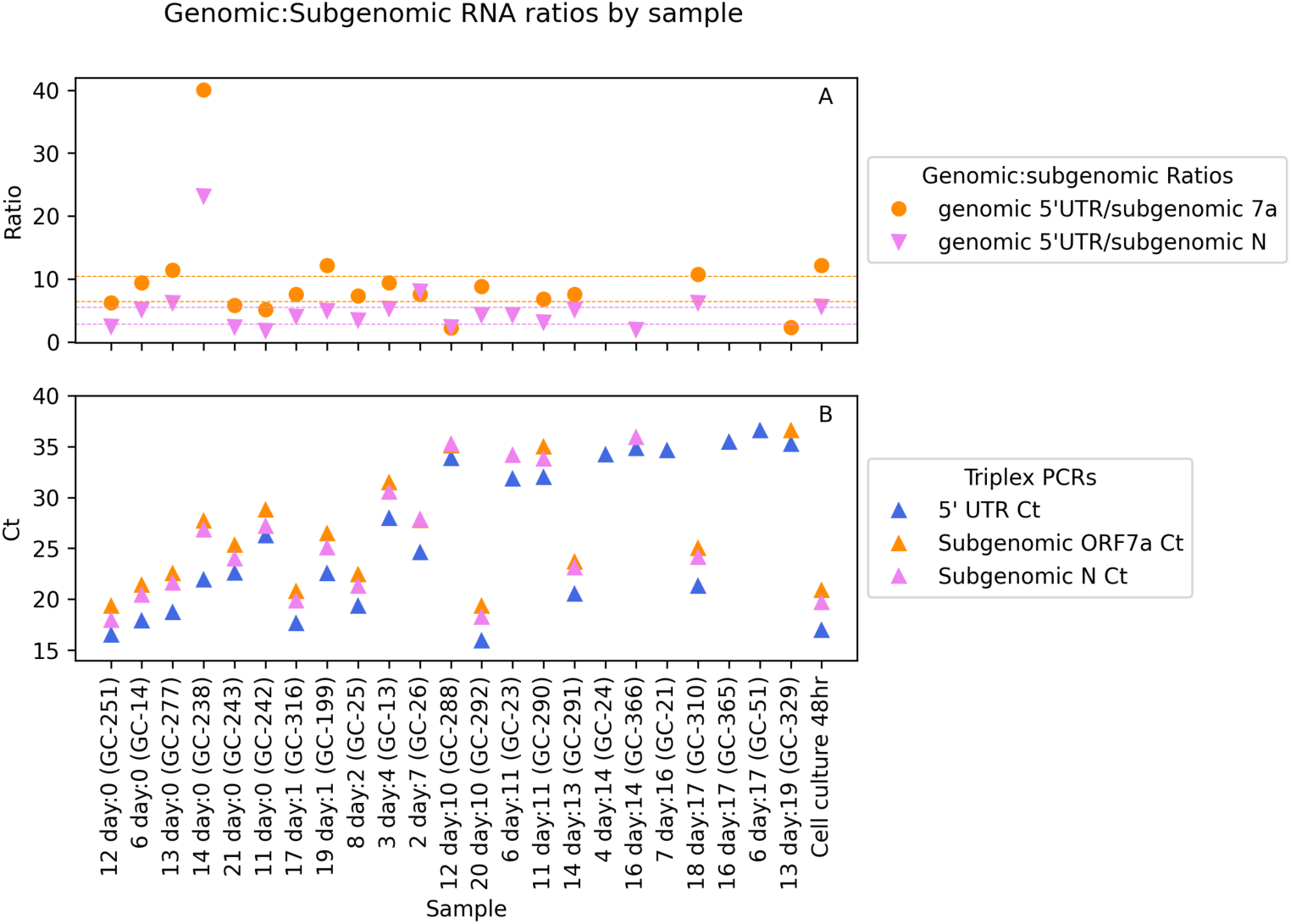
A: The ratio of SARS-CoV-2 genomic RNA molecules to the subgenomic ORF7a and N RNA within the nasopharyngeal swab and cell culture samples. The interquartile ranges of values (6.4-10.4) of the genomic to subgenomic ORF7a RNA ratio are shown as orange dotted lines. The interquartile range (2.9-5.5) of the genomic length to subgenomic N gene RNA ratio are shown as pink dotted lines. B: The triplex PCR Ct’s sorted by the time between symptom onset and swab collection.

For the majority of the nasopharyngeal swab samples, there was, as indicated above, approximately 6-10 fold more genomic than subgenomic ORF7a RNA, and 3-5 fold more full length genomic to subgenomic N gene RNA irrespective of when the swab was collected. Given that we would expect it more likely that replicating virus is present in the swabs collected closer to the onset of symptoms^[5]^, the fact that the ratio of the amount of subgenomic RNA present changes little or not at all between samples collected at symptom onset or two weeks later provides further evidence to our previous observation^[23]^ that the detection of subgenomic RNA should not be considered an indicator of active SARS-CoV-2 replication in nasopharyngeal swabs. The amount of subgenomic RNA is proportional to the total amount of SARS-CoV-2 RNA present in a sample, and therefore the amount of subgenomic RNA could be closely predicted by looking at the Ct of the SARS-CoV-2 diagnostic PCR.

### Detection of positive and negative strand SARS-CoV-2 RNA in diagnostic samples

We next wanted to study whether we could accurately quantify the ratio of positive to negative strand SARS-CoV-2 RNA molecules in clinical samples, as detection of negative strand RNA has been proposed as another surrogate measure of replicating virus^[13]^. To do this, we used the specific probe based PCR assays mentioned above, but instead of making cDNA with random hexamers, we generated strand specific cDNA with the specific forward or reverse sense primers for the negative and positive strand assays respectively (See Material and Methods). During our assay development, we found that the reverse 5’UTR PCR primers designed to specifically detect SARS-CoV-2 genomic RNA, while relative efficient PCR primers on cDNA prepared by random hexamer priming (see above and Materials and Methods), performed very poorly as a cDNA synthesis primers from positive strand templates. This could possibly be explained by the recently reported secondary structure of the 5’UTR of the SARS-CoV-2 genomic RNA which revealed that the location where our reverse 5’UTR primer annealed lay within a highly stable RNA hairpin structure (stem loop 5a), which has been shown to be resistant to RNase I treatment even under denaturing conditions^[27]^. As a result, this assay was not appropriate to study the ratios of positive to negative sense RNA molecules in the clinical samples most likely due to poor reverse primer annealing and subsequent cDNA synthesis. The ORF7a total RNA, ORF7a subgenomic and N subgenomic RNA PCRs however, all performed well and after the initial optimisation performed on dilutions of the RNA from the SARS-CoV-2 positive cell culture, all had efficiencies of close to 95% when tested on serial dilutions of control PCR amplicons (See Material and Methods). From the non-strand specific PCR results above, we knew that the total ORF7a total RNA assay Ct’s were close to those of the 5’UTR PCR, and therefore the majority of targets detected by this assay were likely SARS-CoV-2 genomic-length RNA molecules. Therefore, this PCR could be used to approximate the amount of strand specific SARS-CoV-2 genomic length RNA. From ours and studies performed in other research groups, we knew that the strand specific PCRs were less sensitive than their non-strand specific counterparts likely due to a lower efficiency of the single primer initiated cDNA synthesis as well as the negative strand RNAs being less abundant than positive sense RNAs in SARS-CoV-2 and other coronaviruses^[20,23,28,29]^. Therefore for this experiment, we selected fourteen of the swab samples with the lowest 5’ UTR Ct’s along with the cell culture sample.

The strand-specific ORF7a total PCR assay was able to detect positive sense SARS-CoV-2 genomic length RNA (ORF7a total) in all the fourteen naso-oropharyngeal swabs tested as well as the cell culture sample (Table 3 and Figure 3). Similarly, positive sense N gene subgenomic RNA was detected in all samples, but positive sense ORF7a subgenomic RNA was only detected in eight swab samples and the cell culture sample. Negative sense genomic length ORF7a total and N gene subgenomic RNAs were detected in eleven swab samples, while negative sense ORF7a subgenomic RNA was detected in 9 swabs (Figure 3). The strand specific PCRs were repeatable, although some samples with very high Ct’s >35 for any of the targets tested negative in one of the repeats. This indicated that some of these targets were near the limit of detection of the assays, and that the PCRs were now stochastically amplifying the targets at these very high Ct’s. We therefore excluded sample GC-26 from the ratio calculations below as it had negative sense ORF7a total, subgenomic ORF7a and N gene PCR Cts of 34.7, 38.3 and 35.4 respectively. GC-291 and GC-238 similarly reported a negative sense subgenomic N gene Ct of 36.1 and 34.7 and were excluded from subgenomic N gene strand specific ratio calculations below. Some of our other nasopharyngeal swab samples were nearly exhausted and we were not able to repeat all strand specific assays for all samples (see Table 3).

**Table 3.** Ct values obtained by the strand specific RT-PCRs of 14 SARS-CoV-2 positive nasopharyngeal swabs and the 48hr cell culture supernatant. Individual 1 was used as a negative control

| Individual | Swab Sample ID | Days since onset of symptoms/Initial swab | ORF7a total +ve strand |  | ORF7a total -ve strand |  | ORF7a subgenomic +ve strand |  | ORF7a subgenomic -ve strand |  | N gene subgenomic +ve strand |  | N gene subgenomic -ve strand |  |
| --- | --- | --- | --- | --- | --- | --- | --- | --- | --- | --- | --- | --- | --- | --- |
| 1 | GC-28 | 0* | NEG | NEG | NEG | NEG | NEG | NEG | NEG | NEG | NEG | NEG | NEG | NEG |
| 2 | GC-26 | 7 | 24.3 | 24.6 | 33.2 | 34.7 | 33.2 | 34.1 | NEG | 38.3 | 29.6 | 29.8 | 39.2 | 35.4 |
| 3 | GC-13 | 4 | 35.8 | 35.3 | NEG | NEG | NEG | NEG | NEG | NEG | 34.6 | 33.3 | NEG | NEG |
| 6 | GC-14 | 0 | 20.6 | 20.4 | 27.7 | 28.2 | 25.6 | 24.8 | 31.2 | 31.6 | 24.1 | 24.1 | 27.9 | 27.7 |
| 8 | GC-25 | 2 | 21.3 | 21 | 26.9 | 27.2 | 25.6 | 25.4 | 31 | 31 | 24.2 | 24.1 | 29 | 29.2 |
| 11 | GC-242 | 0 | 27.8 | 28.1 | NEG | NEG | NEG | NEG | 37.5 | NEG | 30.7 | 30.5 | NEG | NEG |
| 12 | GC-251 | 0 | 18 | 17.8 | 26.1 | 27.3 | 22.5 | 22.3 | 33.1 | 32.2 | 20.6 | 20.6 | 28.6 | 28.5 |
| 13 | GC-277 | 0 | 20.4 | 20.5 | 29 | 28.6 | 24 | 25.5 | 31.9 | 33.5 | 22.7 | 23.8 | 29 | 28.4 |
| 14 | GC-238 | 0 | 24 | 24.4 | 33.8 | 33 | NEG | NEG | NEG | NEG | 29.4 | 28.8 | 34.7 | NEG |
| 14 | GC-291 | 13 | 22.8 | 22.6 | 30.5 | 30 | NEG | ND | NEG | ND | 27.8 | ND | 36.1 | ND |
| 17 | GC-316 | 1 | 20.4 | 20.5 | 26.2 | 25.3 | 27.3 | ND | 34.3 | ND | 23.8 | ND | 29.5 | ND |
| 18 | GC-310 | 17 | 23.6 | 23.1 | 29.9 | 29.8 | NEG | ND | NEG | ND | 27.9 | ND | 32.7 | ND |
| 19 | GC-199 | 1 | NEG | 30.9 | NEG | NEG | NEG | 38.1 | NEG | NEG | 27 | 26.9 | NEG | NEG |
| 20 | GC-292 | 10 | 18.8 | 19 | 24.3 | 24.6 | 25.7 | ND | 31.4 | ND | 22.1 | ND | 26.9 | ND |
| 21 | GC-243 | 0 | 24.1 | 24.6 | 32.6 | 30.4 | NEG | NEG | 34.2 | 34.3 | 27.2 | 26.8 | 31.4 | 31.3 |
| SARS-CoV-2 Negative swab pool | Pool of 10 Neg. swabs | NA | NEG | NEG | NEG | NEG | NEG | NEG | NEG | NEG | NEG | NEG | NEG | NEG |
| Cell culture 48Hr |  | NA | 14.6 | 15.1 | 25.5 | 24.7 | 20.9 | 21.2 | 33.2 | 33.5 | 19.5 | 19.4 | 31.6 | 30.4 |

ND: insufficient RNA remaining to perform repeat PCR
\*Date of onset of clinical signs not provided. Number of days since the collection of the first nasopharyngeal swabs is shown.

**Figure 3.**
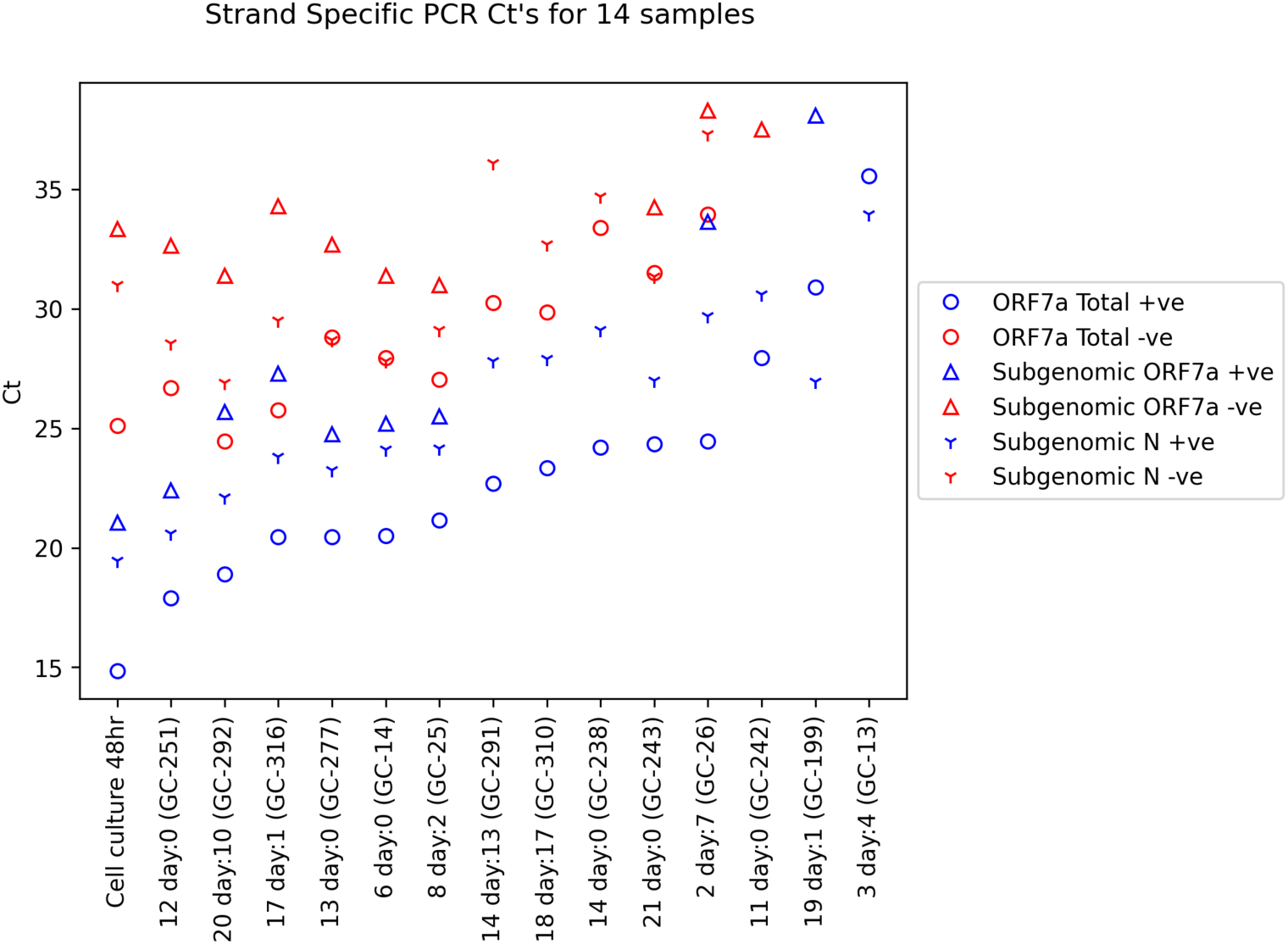
The Ct values of the strand-specific PCRs of the cell culture and the 14 nasopharyngeal swabs with the lowest Ct’s. Samples have been arranged in ascending order based on the Total ORF7a +ve strand PCR. (n=14 data points from 14 samples run twice)

In the samples with a positive sense ORF7a total PCR Ct of 20 or less, all negative and positive sense targets were detected. In samples with a positive sense ORF7a total Ct > 20, detection of one or more strand specific targets became inconsistent, and only one of those samples (GC-26 taken on day 7 from individual 2) was PCR positive for all of the positive and negative strand targets, albeit at very high Ct for the negative sense subgenomic targets. In samples with a positive sense ORF7a total PCR Ct greater than 30, we did not detect any negative sense RNAs. Negative sense RNA could be detected in samples collected up to 13 and 17 days post the onset of symptoms (Figure 3).

Given there appeared to be a relationship between the strand specific assays based on the Ct of the positive sense total ORF7a PCR Ct, we decided to plot the strand specific assays against the Ct from the non-strand specific 5’UTR to see if detection of negative strand targets was a product of the amount of SARS-CoV-2 RNA present in the sample. The strand specific total ORF7a and subgenomic N gene PCR Ct’s were strongly correlated (Spearman correlation coefficients (ρ) of 0.80 to 0.96) with the the 5’ UTR Ct (Figure 4). The Ct of the strand specific subgenomic ORF7a PCRs was less strongly correlated with the Ct of the 5’UTR (positive strand: ρ=0.55; negative strand ρ=0.53), however these two assays always had higher Ct’s than the PCRs for the other targets. Therefore these assays may not have been as accurate in measuring the load of strand specific subgenomic ORF7a RNAs in this higher Ct range where the PCR could behave stochastically. The more abundant RNA molecules showed a close to linear relationship with the 5’UTR PCR, including the positive and negative strand ORF7a total RNA (ρ=0.96 and ρ=0.94) and the positive and negative sense subgenomic N gene RNA (ρ=0.94 and ρ=0.8). The strand specific Ct’s correlated poorly (−0.09 ≤ ρ ≤ 0.64) with the time between symptom onset and swab collection, and there was no obvious pattern descernable from the scatterplots of these two variables (Supplementary Fig. S3).

**Figure 4.**
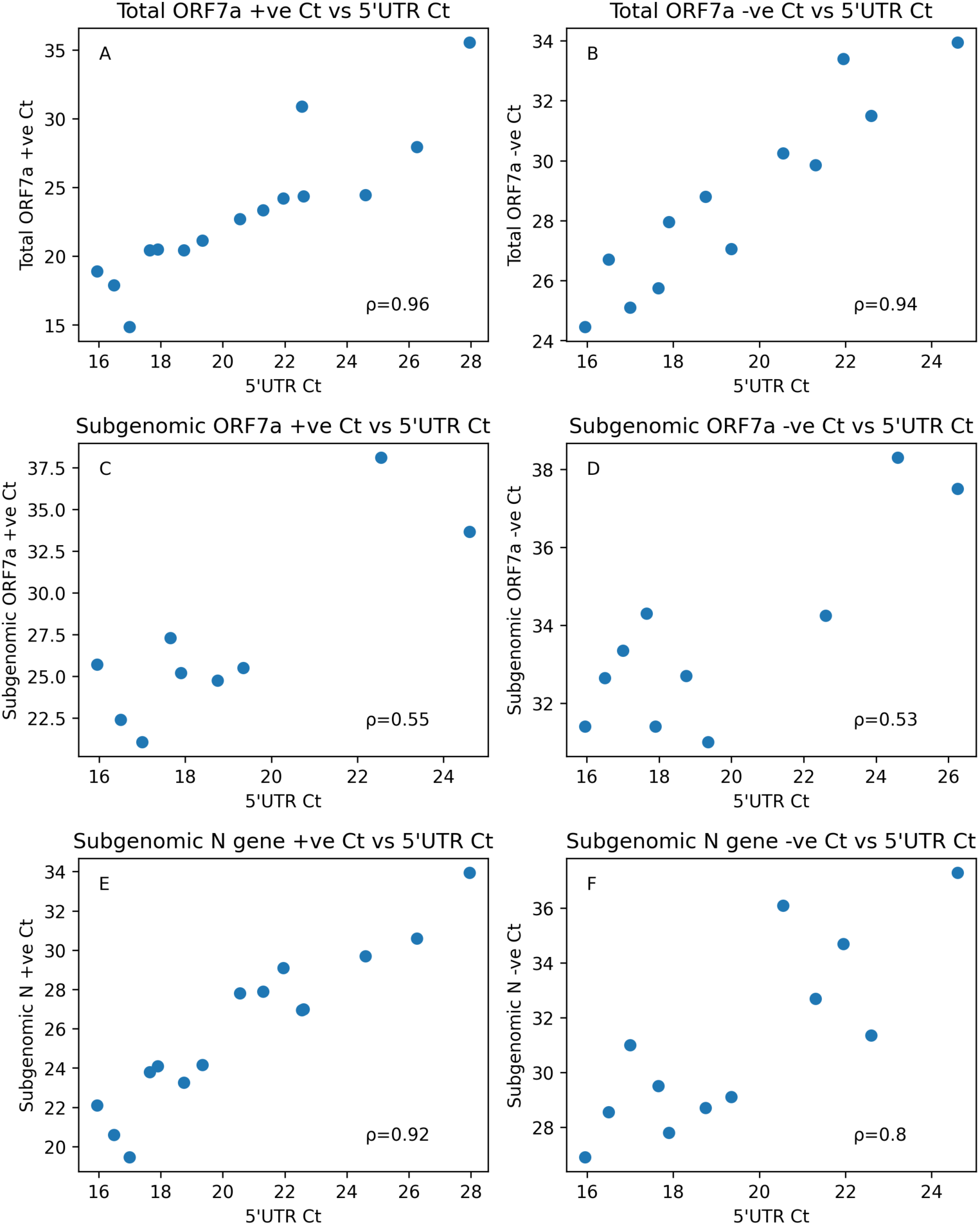
Scatter plots of the Ct of each strand specific PCR against the triplex 5’UTR Ct. For each strand specific PCR, there was a strong correlation between the Ct of the 5’ UTR (Spearman’s ranked correlation coefficient (ρ) shown in each plot). (n=9 to 15 datapoints from 15 samples run twice).

We next looked at the ratios of positive to negative strand RNA for each PCR target. This was calculated by 1.9^(*negative strand PCR Ct− positive strand PCR Ct*)^(1.9 representing 95% PCR efficiency) for each target to determine how much more positive strand relative to negative strand RNA existed for each target. We plotted the ratios for each PCR target against the time between swab collection and symptom onset (Figure 5). The ORF7a total median positive to negative strand RNA ratio was 108.9 times (IQR: 49-191) more positive strand to negative strand RNA across the swabs. The subgenomic ORF7a had a median 71.4 (IQR: 42.5-145.7) times more positive sense than negative sense RNA and the subgenomic N gene had a median 24 (IQR: 21.8-36.4) times more positive sense than negative sense RNA. While the majority of molecules of each RNA target were positive sense, the relative proportion of negative strand SARS-CoV-2 RNA was highest for the subgenomic N gene, followed by the subgenomic ORF7a and then the genomic full length RNA. The subgenomic N gene RNA has also been observed to have the highest relative amount of negative strand RNA in other coronaviruses such as porcine transmissible gastroenteritis virus^[28]^ and mouse hepatitis virus^[29]^. There was low to moderate negative correlation (−0.13 < ρ < -0.76) between the time between swab collection and symptom onset and the positive to negative strand ratios (Figure 5). We did note that some of the swabs collected at the onset of symptoms did have higher proportions of positive strand RNA particularly in the total ORF7a assay. Three swabs, GC-238, GC -251 and GC-277 in particular had positive to negative ratios higher than 200 and therefore above the 75^th^ percentile (191) of the positive to negative ratio for this target. If a sample had a high number of virions present, it is possible that this might be observed as a high positive to negative ORF7a total ratio, and it would be more likely that a sample taken closer to the onset of symptoms would have infectious virus particles present^[5]^. Therefore it might be possible to determine if a sample is more likely infectious by looking at the ratio of positive to negative genomic length RNA. We did not however have access to appropriate biosafety facilities in our laboratory to attempt culture from these swabs to test this hypothesis. Even if this ratio was predictive of infectivity, the test would likely only be useful in samples with a high viral RNA load to ensure the negative strand ORF7a total PCR was operating in a Ct range (ie a Ct<35) where the amplification of the RNA targets was not behaving stochastically. The single cell culture sample had very high positive to negative strand ratios for all three targets. The ORF7a total ratio was 719.8, the subgenomic ORF7a ratio was 2683.3 and the subgenomic N gene ratio was 1658. These ratios were significantly higher than those seen in the nasopharyngeal swabs. We speculate that these ratios were higher in this sample for three reasons. Firstly, unlike the nasopharyngeal swabs, any virions produced in the cell culture would have accumulated in the media and therefore the ORF7a total positive to negative ratio could be elevated as a result. Secondly, this sample was gamma-irradiated prior to leaving the biosecure facility in which it was grown, and this likely would have disproportionately affected double stranded RNA by causing crosslinking between the two strands. As most of the negative strand coronavirus RNA is understood to exist as double stranded molecules^[20,22,29,30]^, this effect would have caused a relative decrease in the amount of negative strand RNA measured by the PCR assays. Thirdly, the sample was clarified by centrifugation which might have removed cells and larger double membrane structures from the media, and therefore removed a relatively higher proportion of the negative compared to positive strand RNA.

**Figure 5.**
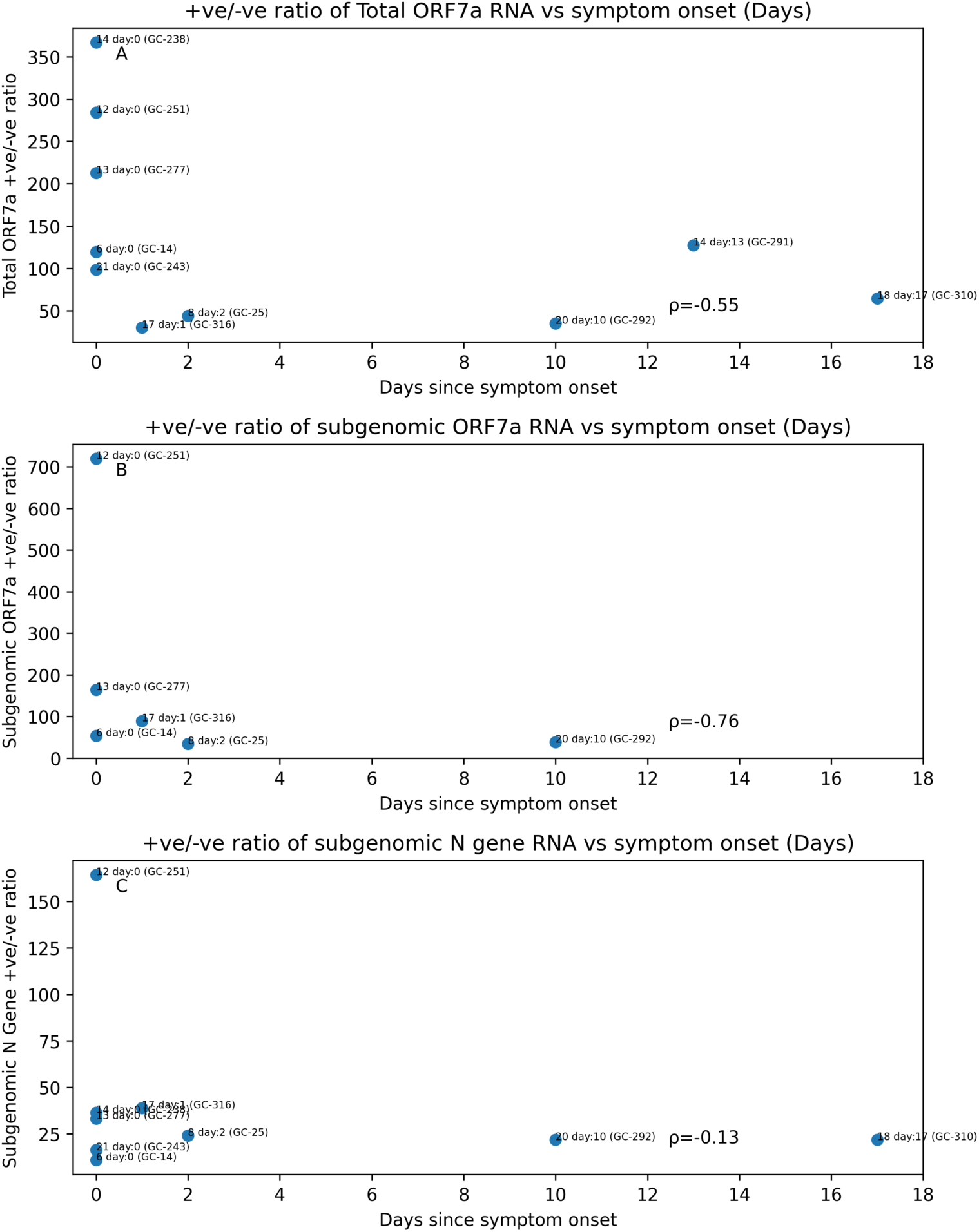
The positive to negative strand ratio for the ORF7a total, subgenomic ORF7a and subgenomic N gene plotted against the time of swab collection since the onset of symptoms. Spearman’s ranked correlation coefficient (ρ) is shown in each plot (n=14 datapoints from 14 samples run twice)

### Detection and abundance of NGS reads mapped to subgenomic RNAs

To further explore the pattern of subgenomic RNAs in the naso-oropharyngeal swab samples, we created two Ampliseq primer mini-panels using 11 reverse primers within each of the ten potential SARS-CoV-2 canonical subgenomic RNAs and the 5’ UTR region together with two different forward primers within the 5’ leader sequence of all the SARS-CoV-2 RNAs. These primers were a subset of the Ampliseq primers from the ThermoFisher Ion AmpliSeq SARS-CoV-2 Research Panel which we had previously used to detect subgenomic RNAs^[23]^. The rationale for doing so was to essentially create a multiplex PCR to quantitate the relative abundance of genomic and subgenomic RNAs within each sample. We found that despite these two mini-panels differing only in their forward primer, the amount of amplification detectable in the same SARS-CoV-2 positive swab samples differed greatly with an average of 901,000 reads generated with mini-panel 2 after 31 cycles, and an average of 21,000 reads generated by mini-panel 1 after 31 cycles of PCR amplification. The majority (80%+) of the reads from mini-panel 1 were not mapped to the SARS-CoV-2 genomic length or subgenomic RNAs (see Source Data). Given that the forward primer was the only difference between the two mini-panels, this primer was the likely reason for the lower amplification in mini-panel 1. We subsequently identified during the course of full genome sequencing (see below), that there was a mismatch between this primer and the SARS-CoV-2 genome sequence of several, perhaps all, of the samples we had used in this study, and this likely resulted in this primer annealing poorly during the Ampliseq PCR across the 5’UTR and subgenomic amplicons. As a result, we found that we needed to use 31 cycles of PCR amplification with this panel to obtain an average of 459 reads mapped to all of the amplicon targets (see Source Data).

With 31 PCR cycles, all canonical subgenomic RNAs except for ORF7b and ORF10 were detected by mini-panel 1. Subgenomic N gene was the most abundant subgenomic molecule, followed by ORF7a (Figure 6). These two molecules were also the most abundant in the cell culture sample (See Source Data). Together, these two molecules made up between 50-95% of all the subgenomic RNA within the samples. The other subgenomic RNAs were present, but at lower abundances. A similar pattern was also seen at 21 PCR cycles although with relative lesser amplification of the lower abundant subgenomic targets by that stage of the PCR reaction (Supplementary Fig. S4). Making cDNA with the panel primers resulted on average in a similar number of reads at 21 cycles as the random hexamer cDNA (316 vs 339) (See Source Data). Again, subgenomic ORF7a and N were the most abundant amplicons irrespective of the method of cDNA creation.

**Figure 6.**
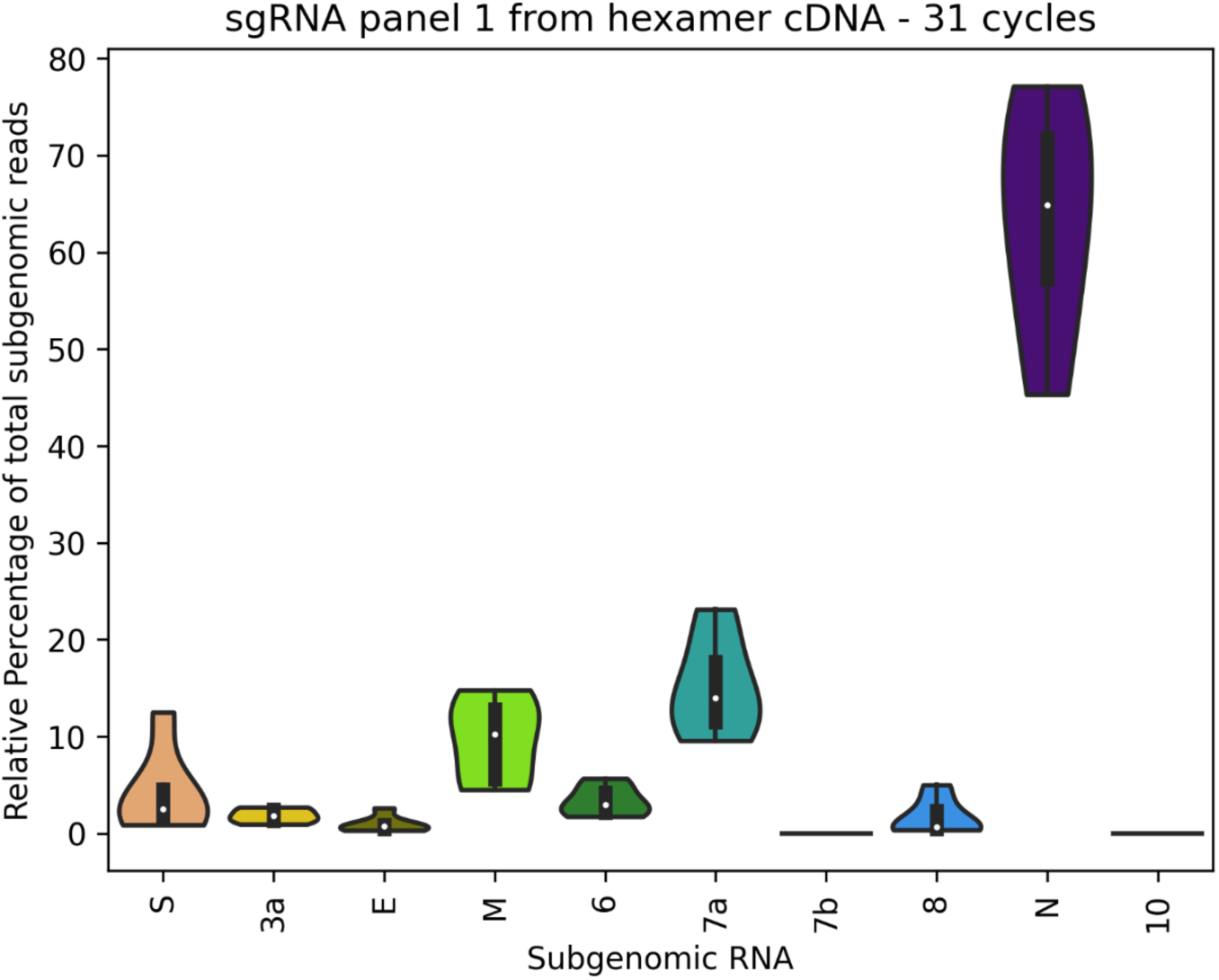
Relative percentage of each subgenomic RNA of all reads mapped to the SARS-CoV-2 subgenomic RNA for the Ampliseq mini-panel 1 in naso-oropharyngeal swab samples with 31 PCR amplification cycles. Subgenomic ORF7a and N gene RNAs were consistently the two most abundant subgenomic RNAs present in the samples. Subgenomic ORF7b and ORF10 were not detected in any sample. (n=12 from 12 biological samples run once)

In mini-panel 2, the forward primer was much more efficient than the mini-panel 1 forward primer, and we saw much more amplification across all targets. The 5’ UTR amplicon was particularly efficient, and even by 21 cycles of amplification (Figure 7), the number of reads mapped to this amplicon was significantly higher than all other amplicons (see Source Data). By 31 cycles (Supplementary Fig. S5) we could see that the relative number of reads mapped to the subgenomic RNAs was reduced due to this single amplicon crowding them out on the sequencer chip. Despite this, mini-panel 2 identified a similar pattern in the relative abundance of the subgenomic RNA’s in both the 21 and 31 cycles of amplification, with the subgenomic N gene and subgenomic ORF7a RNAs being the two most abundant subgenomic RNAs across the swab samples (making up between 82.3-97.5% of all subgenomic reads at 21 cycles, and 88.9-100% at 31 cycles), and the other canonical subgenomic RNAs being expressed at much lower levels (other than ORF7b and ORF 10 which were not detected at all) (Figure 7 and Supplementary Fig. S5). Subgenomic N and ORF7a RNAs were also the most abundant SARS-CoV-2 subgenomic RNA molecules in the cell culture sample (See Source Data).

**Figure 7.**
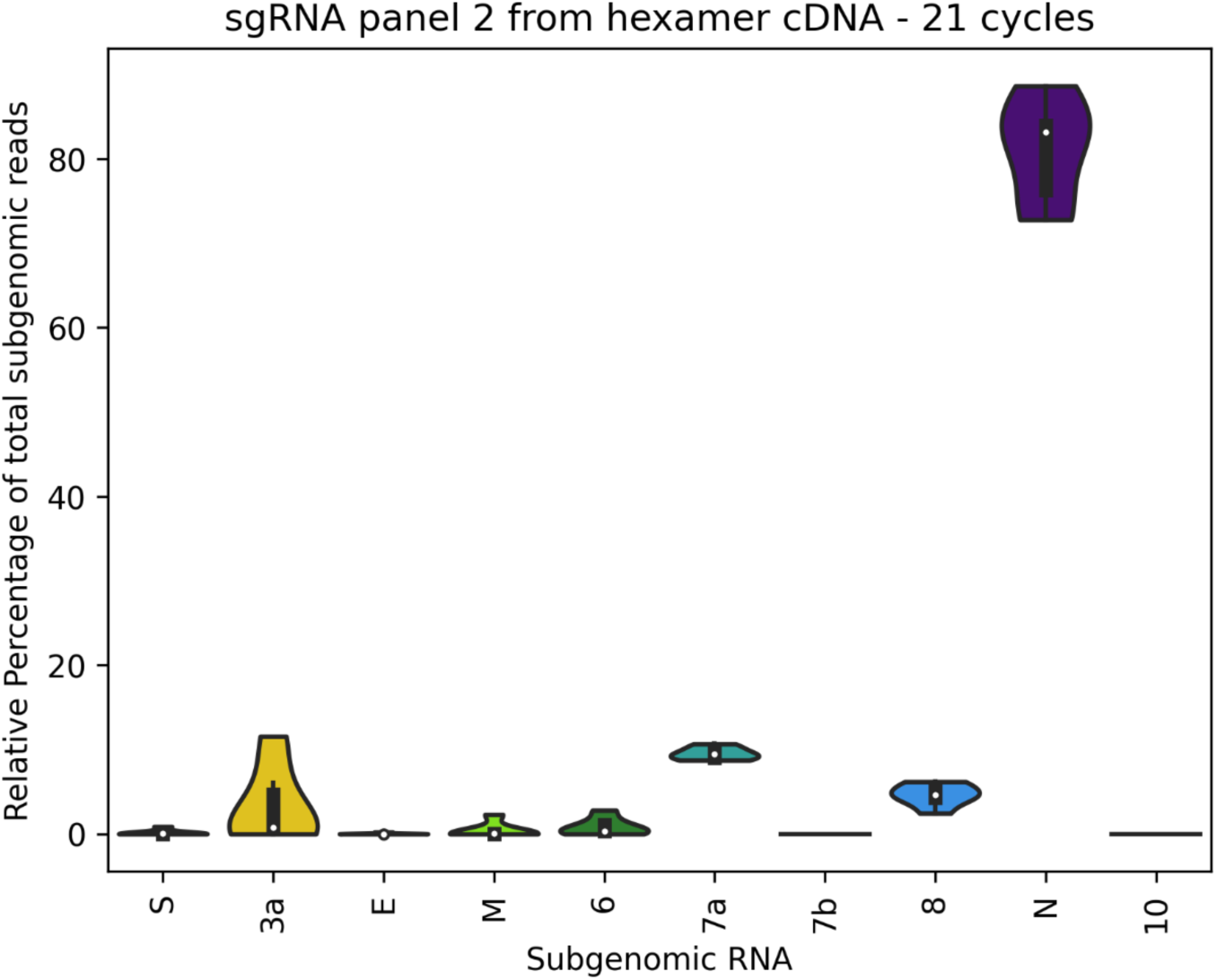
Relative percentage of reads mapped to each subgenomic RNA using the Ampliseq mini-panel 2, cDNA synthesis with random hexamers and 21 PCR amplification cycles. (n=6 data points from 6 biological samples run once)

Mini-panel 2 was able to detect a low number of reads belonging to potential non-canonical subgenomic RNAs which could initiate expression of the proteins of ORF 9b^[2,31,32]^ and also ORF 7b. The ORF9b subgenomic RNA contained the 69 nucleotides of the leader joined to nucleotide 28284, which is 24 nucleotides downstream of the regular N gene TRS (based on the nucleotide numbering of SARS-CoV-2 Wuhan-Hu-1 NC_045512.2). The non-canonical ORF 7b subgenomic RNA, here designated subgenomic RNA 7b*, contained the 69 nucleotides of the leader sequenced joined to nucleotide position 27674, 26 nucleotides downstream of the canonical 7b TRS site. The ability to detect such reads appeared to be related to the number of sequencing reads, and therefore these particular subgenomic reads were seen more frequently when we used 31 cycles of PCR amplification. The number of subgenomic ORF 9b reads made up only approximately 0.03-0.11% of the total subgenomic N gene reads and the number of subgenomic ORF 7b was equal to only 0.13-0.25% of the number of subgenomic ORF 7a reads (See Source Data). Interestingly, there was some minor heterogeneity to these non-canonical subgenomic RNAs in the occasional read in some samples such as GC-14 (using 21 PCR cycles, see Source Data) which showed the leader joined to sequence upstream of the 7b AUG at position 27602 and also the 9b subgenomic RNA had some minor heterogeneity, for example for sample GC-251 (using the full panel (see below) and 21 cycles, see source data). Close inspection of reads generated in our previous study^[23]^ also supported this minor heterogeneity, for example GC-277 and GC-26 which showed the leader joined to sequences at position 27678 for the 7b* subgenomic RNA and some heterogeneity around the 9b site, respectively (data not shown, raw data available in SRA Accession PRJNA636225).

### SARS-CoV-2 Sequencing and subgenomic analysis using the full Ampliseq panel

We had previously analysed the genome sequence of the SARS-CoV-2 viruses in samples from individuals 2 to 8 which were from the first wave of COVID-19 infections in Victoria in March-May 2020^[23]^, but had not sequenced the later samples from the second wave between July and September 2020 included in this study. The samples from the ten individuals from this period (individuals 11 to 21) were all community acquired infections and likely acquired the infection from several different sources according to the information provided by the local COVID-19 outbreak contract tracing team. Using the full SARS-CoV-2 Ampliseq Panel, we obtained the near complete SARS-CoV-2 genomic sequence (nucleotide positions 42 to 29,842) from the swab samples collected from individual 17 day 1 (GC-316) and from four other individuals who were linked to the same case cluster by the contact tracing team - individuals 12 day 0 (GC-251), individual 14 day 13 (GC-291), individual 20 day 10 (GC-292) and individual 21 day 0 (GC-243) (GenBank accessions: MZ410617-MZ410621). We also obtained partial SARS-CoV-2 sequence from swabs from two cases linked to this cluster – GC-277 (individual 13 day 0, GenBank accession: MZ410622) and GC-238 (individual 14 day 0, NCBI SRA: SRR14836407). The sequences from individual 17, 12, 14 and 21 were identical to each other, although individual 17 was not linked to the same case cluster as individuals 12, 14 and 21. These four sequences were identical to a sequence reported from Victoria from early August 2020 (SARS-CoV-2/human/AUS/VIC17057/2020 GenBank accession: MW321043). The partial sequences from individual 13 was also identical to MW321043 over the genome regions we obtained. All the individuals from the case cluster were tested for SARS-CoV-2 in the first part of August whereas individual 17 may have picked up this virus 2-3 weeks later further along the community transmission chain.

The sequence from individual 20, who also belonged to the case cluster, differed in two nucleotides from MW321043 and the other SARS-CoV-2 sequences obtained from the other individuals in the cluster. One change at nucleotide 22993 (based on the nucleotide numbering of SARS-CoV-2 Wuhan-Hu-1 NC_045512.2) was a change from a C to T without altering the amino acid sequence of the spike protein. The second change, G to A, was at nucleotide 28884 located within the N protein (amino acid 203) and changed an arginine to a glutamine in the nucleoprotein sequence. This would have also resulted in a lysine to glutamic acid in the ORF9c protein (amino acid 50)^[33]^, and altered the non-canonical TRS of the proposed ORF N*^[33]^ which was present in the other SARS-CoV-2 sequences presented here. Searching for sequences from the second wave in Victoria, Australia with BLASTN on GISAID and GenBank’s nucleotide database, we found that there was only one virus sequenced in the local epidemic with this change in the nucleoprotein during the initial epidemic molecular tracing (SARS-CoV-2/human/AUS/VIC8533/2020, GenBank accession: MW153442) collected in early August 2020. The characteristics for MW153442 matched those of our individual 20, and subsequent follow up of laboratory records revealed that individual 20 was in fact tested for SARS-CoV-2 in early August, and the swab sample sent to the state reference laboratory for sequencing. Given that no other virus sequences with this change were identified in the local epidemic, this variant may have only arisen in a single individual during the outbreak in this cluster, but was not able to, or did not have the opportunity to transmit and propagate significantly in the general population afterwards. A MegaBLAST search in NCBI’s Nucleotide database revealed that the same amino acid change in the nucleoprotein has been observed in other countries indicating that SARS-CoV-2 virus has spontaneously made this change multiple times during the global epidemic. Interestingly, in the SARS-CoV-2 sequencing from the swab samples GC-251 and GC-243 from individuals 12 day 0 and 21 day 0, the forward primer of the first 5’UTR amplicon of pool 1 of the Ampliseq panel did not work well. As a result, several reads were created by linear amplification from the reverse primers binding to positive sense cDNA which were able extended beyond where the forward primer would sit. This revealed a change of a C to a T at nucleotide position 40 within the leader sequence, which is within the 3’ end of where the corresponding 5’UTR forward primer would anneal. This change is most likely the reason the forward primer was unable to anneal and amplify in these two samples. Given that all the other SARS-CoV-2 sequences we generated in this study, except for that of individual 20, were identical to the SARS-CoV-2 genomes from individuals 12 and 21, it is entirely possible that those SARS-CoV-2 genomes also had the same C to T change at position 40. However in those samples a forward primer likely did manage to eventually anneal during the amplification PCR of these samples resulting in the production of the expected amplicon. If this nucleotide change was present in the samples, then this change would lie in the 3’end of the forward primer of mini-panel 1 which used the same forward primer as pool 1. This could explain why mini-panel 1 amplified the 5’UTR and subgenomic RNAs inefficiently compared to mini-panel 2. For mini-panel 2, the 3’ end of the primer was at nucleotide position 52, and therefore this particular nucleotide substitution would have sat within the middle of the primer which is less critical to primer annealing and extension when compared to the 3’ end^[34]^. There was also a nucleotide change of a C to a T at position 241 in all seven SARS-CoV-2 genomes sequenced in this study. This change sat within the middle of the annealing site of the reverse 5’UTR primer of both mini-panels. Its effect on the primer’s ability to anneal was likely minor, as the 5’ UTR was the most efficient amplicon of mini-panel 2.

By looking for reads with the 5’ UTR leader sequence joined to 3’ ORFs at known TRS sequences, we could identify between 161 to 142999 reads coming from subgenomic RNA in the full Ampliseq panels (See Source Data). Consistent with the mini-panels, reads mapping to subgenomic ORF7a and N gene RNAs were the most abundant, accounting for 37.3 to 61.2% of all subgenomic reads (Supplementary Fig. S6). The full panel was also effective at amplifying ORF3a and ORF6 subgenomic RNAs, and therefore it estimated the relative abundance of these two RNAs higher than either of the two mini-panels. The remaining canonical subgenomic RNAs were also detected (except for ORF7b and ORF10) but made up a much smaller fraction of the total subgenomic reads (11-23% of subgenomic reads). Like mini-panel 2, the full panel was also able to detect a low number of some non-canonical ORF 9b subgenomic RNAs (0-0.65% of mapped subgenomic reads). Sample GC-251 contained a TRS joining variant where the 69nt of the leader was joined to nucleotide 28278, 6nt earlier than ORF 9b subgenomic reads seen with the mini-panel 2 in the samples GC-277, GC-316, GC-291 and GC-292. No non-canonical subgenomic ORF 7b* were detected by the full panel, however subgenomic RNAs which would initiate ORF N*^[33]^ were detected by the full panel (0.27-1.83% of mapped subgenomic reads) (See Source Data). These reads joined the leader nucleotides 1-69 to the nucleotide 28882 at a TRS sequence which was created by a triple nucleotide polymorphism GGG→AAC at nucleotides 28881-2883 which arose in the B.1.1 lineage early in 2020 ^[35,36]^.

Both samples GC-243 and GC-251 reported much fewer subgenomic reads compared to the other swab samples consistent with the nucleotide change in the annealing site of the 5’UTR forward primer sequence resulting in poor primer binding and amplification. In contrast, Sample GC-277 also had reduced 5’UTR amplification using the full panel compared to other samples, but had good amplification of many of the subgenomic targets. Why this occurred is unclear, but perhaps the 5’UTR of the genomic RNA renatured due to its secondary structure^[27]^ during cDNA synthesis in this particular sample.

### Exploring the quality of cellular RNA and mRNA in diagnostic naso-oropharyngeal swabs

As we were looking specifically at RNA intermediates generated during SARS-CoV-2 replication in the oro-nasal mucosa, we wanted to understand whether we could measure how well a swab sample had been collected and transported by looking at host cellular RNA and whether that had any effect on the detection of SARS-CoV-2 RNA. Using a bioanalyzer, we measured the quantity of RNA and the RNA integrity number (RIN) of the extracted nucleic acids from twenty five of the nasopharyngeal swabs, the cell culture sample and one pool of ten SARS-CoV-2 negative nasopharyngeal swabs (see Source Data). The RNA quantity of the nasopharyngeal swabs and swab pools was highly variable (40-2167pg/µl). For the SARS-CoV-2 positive nasopharyngeal swabs, the Ct of the triplex SARS-CoV-2 5’UTR correlated poorly with the amount of RNA present within a swab sample (ρ=0.42, Kendall Rank Correlation=0.3) (Supplementary Fig. S7 and Source Data).

Of the twenty four individual SARS-CoV-2 positive nasopharygeal swabs, the bioanalyzer was not able to calculate a RIN score for eight samples, and reported a RIN of <3 for a further 12 indicating that the cellular RNA of the swab samples was degraded. This may have been due to the infection itself resulting in cellular damage and suppressing normal cellular functions, and could also be partly due to the time taken for transportation and handling at the diagnostic laboratories before arriving at our lab. However, a nasopharyngeal swab, particularly from an individual with a respiratory infection, will likely consists of a mixture of living, dying and dead cells along with cellular debris and mucus, and therefore even a freshly collected swab sample processed immediately is probably going to show some degree of RNA degradation. Only four of the SARS-CoV-2 swab samples we used in this study posessed a RIN of above 3. The bioanalyzer was also unable to provide a RIN value for the pool of 10 SARS-CoV-2 negative swabs and reported a RIN of <3 for the single SARS-CoV-2 negative swab from individual 1 (See Source Data).

We plotted the Ct of the triplex PCR assays against the RIN by grouping samples with a RIN less than 3, a RIN between 3 and 6 and a RIN greater than 6 and saw no clear relationship between the RIN and the SARS-CoV-2 Ct (Supplementary Fig. S8). We did note that that the samples with the lowest 5’UTR Ct’s were in the low RIN group which may support the idea that the virus infection may be contributing to the degradation of the cellular RNA in some samples. Both genomic length and subgenomic SARS-CoV-2 RNA were readily detectable in samples in all three RIN groups, including the samples with a low RIN. This suggests that the rate of degradation of the SARS-CoV-2 RNA was very different, and likely slower than that of cellular RNA in clinical samples. This would be consistent with the SARS-CoV-2 RNA molecules being better protected than cellular RNA from RNases by membrane structures^[23]^. Full length genomic RNA could be protected from RNase activity if packaged in virions, and the replicative forms of full-length and subgenomic RNAs protected by the double membrane structures seen in the cytoplasm of cells infected with SARS-CoV-2 and other coronaviruses^[18,20,21]^. Given the poor quality of cellular RNA, it is likely that any unprotected SARS-CoV-2 RNA (eg. positive sense RNA used for protein translation) would also be readily degraded in clinical samples. Therefore the majority of the SARS-CoV-2 RNA being detected in typical nasopharyngeal swab samples most likely comes from the RNAs protected within virions or double membrane structures.

## Discussion

In this study we developed and used new sensitive probe based PCR assays and Ampliseq panels to further explore the pattern and positive or negative strand specificity of SARS-CoV-2 genomic and subgenomic RNAs in diagnostic swab samples. The triplex probe PCR performed as well, if not slightly better than running the three PCRs separately, and therefore was a useful tool to simultaneously quantitate the relative amount of genomic length SARS-CoV-2 RNA as well as the two most abundant subgenomic RNA molecules, the subgenomic RNAs for ORF7a and N. With this assay, we observed that the amount of subgenomic ORF7a and N gene RNA was highly correlated to the amount of full length SARS-CoV-2 RNA present in a sample, and therefore the presence of subgenomic RNA in a sample could readily be predicted from the Ct of a diagnostic PCR targeting SARS-CoV-2 genomic length or total SARS-CoV-2 RNA. We also observed that the ratio of subgenomic RNAs was more or less constant irrespective of the time between symptom onset and swab collection. We could detect SARS-CoV-2 subgenomic RNAs up to nineteen days after the onset of symptoms likely due to the fact that these RNAs are protected from degradation by double membrane structures generated during earlier SARS-CoV-2 replication, and are therefore relatively stable^[21,23]^. These findings support our earlier study^[23]^ and also agree with other investigators who have also observed that subgenomic RNAs decline linearly along with the total number of SARS-CoV-2 RNAs^[9,25]^, and therefore detection of subgenomic RNAs is not a marker for current replication of SARS-CoV-2.

We were able to successfully adapt our subgenomic ORF7a and N gene PCR assays and the ORF7a total RNA assay into RNA strand specific PCRs to study the relative amounts of positive and negative sense RNAs in nasopharyngeal swabs. We were able to detect negative sense SARS-CoV-2 RNA in samples collected up to seventeen days after the onset of symptoms. With the more abundant RNA molecules, the positive and negative sense total ORF7a RNA and the positive and negative sense subgenomic N gene RNA, we observed a strong correlation with the total amount of genomic length RNA measured by the 5’ UTR assay. With the subgenomic ORF7a strand specific PCRs, we did not observe as strong a relationship with the Ct of this 5’UTR PCR. The subgenomic ORF7a was the least abundant RNA out of those which we measured, and therefore these molecules, particularly the negative sense RNA, were often being measured in the Ct range above 30. In this range the PCR was most likely beginning to operate outside its detection range, and this could explain why we did not see a stronger positive correlation between the subgenomic ORF7a RNA molecules and the total amount of genomic length SARS-CoV-2 RNA in the sample. Given that the most abundant strand specific SARS-CoV-2 genomic length RNA and subgenomic N gene RNA are positively correlated with the total amount of genomic length SARS-CoV-2 RNA, the detection of negative strand RNA is no more likely a predictive marker of active replication than the Ct of PCRs measuring the total amount of SARS-CoV-2 RNA in a swab sample. The fact that negative strand RNAs are protected by double membrane structures^[19,21,23]^ means they can likely persist in cells for some time after replication has concluded.

We did note that in some samples taken at the onset of symptoms, there was an increased ratio of positive to negative strand genomic-length RNA relative to the samples collected later in the clinical course of infection. It would be interesting to investigate in the future whether this increase in positive sense RNA was attributable to virions being present in these samples, as we did not have access to the necessary culture facilities for this study to check for infectious virions. If the increased ratio of positive sense RNA does correlate with the presence of virions, this could make a useful research tool for studying coronavirus infections. However it would be difficult to use this technique as part of mass testing for SARS-CoV-2 as the protocol requires several operator interventions to add primers separately for the strand specific synthesis of cDNA then PCR.

The custom ampliseq panels we designed found that the N gene and ORF7a subgenomic RNAs were the most abundant subgenomic RNAs produced by SARS-CoV-2 during the course of infection consistent with our previous study’s findings using the full SARS-CoV-2 Ampliseq panel^[23]^, and we were again able to detect all canonical subgenomic RNAs except ORF7b and ORF10. Mini-panel 2 also detected reads belonging several non-canonical subgenomic RNAs including non-canonical ORF7b RNAs which could have been used to translate the ORF7b protein. We only saw these reads in samples where we were able to sequence very high numbers of reads from the mini-panel PCRs. This may indicate that SARS-CoV-2 may not be reliant on the canonical TRS for production of the ORF7b protein. The ORF7b subgenomic RNAs were relatively rare and produced at levels lower than that of subgenomic S, which was the canonical subgenomic RNAs with the lowest abundance as measured by the mini and full Ampliseq panels. In future, we would want to modify the forward primer of mini-panel 1 to better prime with mutation changes which have occurred in the SARS-CoV-2 genome since this primer was originally designed in early 2020, and we would likely want to incorporate another primer to detect the subgenomic N* RNA^[33,35]^ which the full Ampliseq panel was able to detect and is present in more recent variants of SARS-CoV-2.

All together, we have developed novel methods to explore the subgenomic RNAs produced by SARS-CoV-2 in vivo and in cell culture. The amount of subgenomic RNA and negative strand RNA appears to be directly related to the total amount of SARS-CoV-2 RNA present within a sample, and therefore neither measure provides additional information in regards to whether a sample contains infectious virions beyond what can be already interpreted from the Ct of SARS-CoV-2 diagnostic PCRs. Studies of SARS-CoV-2 in swabs from patients and in cell culture have shown that the probability of successfully culturing SARS-CoV-2 is related to the load of viral RNA in a sample^[5,10,11]^ and therefore in our opinion this remains the simplest and most reliable molecular measure for predicting possible SARS-CoV-2 infectivity of an individual. The observed relative increase of positive strand to negative strand RNA and whether that may indicate the presence of virions in samples taken at the onset of symptoms is something worth exploring in the future, particularly on swab samples where it is known if culturable SARS-CoV-2 virus is present or not.

## Methods

### Sample details, collection and storage

Samples included in this study were selected from combined nasopharyngeal and oropharyngeal swabs collected as part of ongoing public health surveillance for SARS-CoV-2 in Victoria, Australia from January to August 2020. SARS-CoV-2 positive and negative swabs were identified by PCR at two diagnostic laboratories, and the remaining swab and media were transported to the Geelong Centre for Emerging Infectious Diseases (GCEID) laboratory. A subset of 24 positive swabs from 16 individuals, including 7 from our previous study^[23]^, were selected for this study (Table 1). Individuals were selected on the basis of 1) they had repeated swabs taken on multiple days, and/or 2) their initial swab had a Ct below 30. The swabs came from individuals in the community, and most were not known to be linked to each other except for individuals 11, 12, 13, 14, 20 and 21 which were identified by the local contact tracing team as belonging to a known larger cluster of cases. Eleven SARS-CoV-2 negative combined nasopharyngeal and oropharyngeal swabs were selected from samples identified as negative by PCR during the same period. Ten of these samples were pooled into a single pool of 10 swabs, while one (individual 1 from our previous study^[23]^) was used as an individual negative PCR control.

Basic patient details including age group, gender, epidemic wave during which swab was collected, swab collection in days post symptom onset, summary clinical signs and hospitalization or not were collected from laboratory submission forms or medical records. Summary details of the samples included are shown in Table 1. The study complied with all relevant guidelines and ethical regulations and has been approved by the Barwon Health Human Research Ethics Committee (Ref HREC 20/56) and all participants gave their informed consent.

A SARS-CoV-2 positive cell culture supernatant was included as a positive control and for comparison to our diagnostic swab samples. This sample consisted of the supernatant from a 48 hour third passage of SARS-CoV-2 virus in Vero E6 cells and kindly supplied by the Australian Centre for Disease Preparedness. The cultured virus isolate is designated VIC-01-059 and isolated from a SARS-CoV-2 positive individual in Victoria, Australia in 2020. The culture supernatant was clarified at 4000g for 10 min before being gamma irradiated and transferred to the GCEID laboratory.

### Nucleic acid extraction and cDNA synthesis

Nucleic acid extraction was performed on 50 µl of the collection media from the combined nasopharyngeal/oropharyngeal swab samples using the MagMax 96 Viral RNA Isolation kit (Thermofisher) on a Kingfisher Flex extraction robot (Thermofisher). cDNA synthesis was performed by heating extracted nucleic acids at 65°C for 5 minutes and rapid cooling on ice before cDNA synthesis using SuperScript™ VILO™ Master Mix (Thermofisher Scientific, Victoria, Australia) as per manufacturers’ instructions ^[37,38]^ and described previously^[23]^. The nucleic acids from the cell culture positive control sample was extracted using the Qiagen Viral RNA mini kit (Qiagen, Victoria, Australia) as per the manufacturer’s instructions.

### Development of SARS-CoV-2 genomic and subgenomic probe based real-time PCR assays

We had previously developed and reported SYBR/Syto 9 based PCR assays to detect genomic and subgenomic SARS-CoV-2 RNA in clinical samples^[23]^. These assays, however, had limitations in their sensitivity and needed to be run individually as they all used a single fluorescent reporter SYBR/Syto 9 in the real-time PCR reaction. To improve the sensitivity of these assays, we designed four probe-based PCR assays, three of which were similar to our previous SYBR/Syto 9 based assays, and each amplified either the 5’ UTR genomic length RNA, the ORF7a subgenomic RNA only, or the total (genomic and subgenomic) ORF7a RNA, respectively, and a fourth PCR targeting the N gene subgenomic RNA only. Primer and probe sequences were designed based on the sequence of SARS-CoV-2 Wuhan-Hu-1 (NC_045512.2) and are provided in Supplementary Table S3. We tested several forward primers within the leader sequence and found the optimum primer to be one which annealed in the leader sequence of the SARS-CoV-2 genome between nucleotide positions 45 and 66, while the probe and reverse primer annealed in the 5’ UTR downstream of the leader sequence which is only present in the genomic length SARS-CoV-2 RNA. The subgenomic ORF7a and subgenomic N RNA specific assays utilized the same forward SARS-CoV-2 leader primer as the 5’ UTR assay mentioned above, while their probes and reverse primers annealed within their respective genes. They would therefore only detect the subgenomic RNAs where the virus had joined the leader immediately upstream of these genes. The ORF7a total PCR utilized the same probe and reverse primer as the ORF7a subgenomic PCR but used a different forward primer annealing within the ORF7a gene. This PCR would therefore detect genomic length RNA as well as several subgenomic RNA molecules, specifically those used to transcribe the Spike, ORF3a, Envelope, Membrane, ORF6 and ORF7a subgenomic RNAs. The three probes each had a different fluorophore attached, with the intention to be able to combine the assays into a single reaction.

Each of these assays were initially established as single/individual PCRs. For these single assays, 2ul of prepared cDNA from the samples were added to a PCR mastermix comprising 1x Brilliant Multiplex qPCR mastermix (Agilent Technologies, California, USA), 1µM of both the forward and reverse primer, 0.2 µM probe and nuclease free water to a total reaction volume of 10 µl. The PCR reaction was then performed in a Quantstudio 6 real-time PCR machine (Thermofisher, Mulgrave, VIC, Australia). Known SARS-CoV-2 positive and negative swab samples and the SARS-CoV-2 cell culture sample were used to initially optimize the PCR temperature cycling conditions, with the optimum cycling temperatures determined to be 95°C for 10 min, then 40 cycles of 95°C for 3 sec, 58°C for 30 sec and 64°C for 30 sec.

After the successful amplification from known positive clinical samples, the resulting PCR product was visualized on a 2% agarose gel (2% Size Select E-gel, Thermofisher) to confirm a product of the expected size was produced. The amplicon was then extracted from the gel and sequenced using the 3.1 Big Dye Terminator PCR sequencing reaction and sequenced on a Hitachi 3500 genetic sequencer (Thermofisher) to confirm that the amplicons were indeed the expected genomic and subgenomic targets. The gel purified amplicons were quantitated on a QiaXpert spectrophotometer (Qiagen, Hilden, Germany) and used as standards in serial dilution in the subsequent qPCR reactions, and in determining the efficiency of the PCR reactions. The PCR efficiency, estimated to be 95% for these assays, along with the difference in Ct values between the genomic 5’UTR and the subgenomic ORF7a and N assays was used to calculate the ratio of genomic to subgenomic RNA within the samples.

To determine if we could multiplex the individual assays and develop a single test which could both quantitate genomic length and subgenomic RNA in a SARS-CoV-2 positive sample, we initially combined the 5’ UTR and the ORF7a subgenomic assays into a duplex real-time PCR. This was performed by combining 1x Brilliant Multiplex qPCR mastermix (Agilent), 0.9µM of the shared SARS-CoV-2 leader forward primer, 0.9 µM of each of the 5’UTR and ORF7a reverse primers, 0.18 µM of each of the 5’UTR and ORF7a probes, 2ul of sample/control cDNA and nuclease free water to a final volume of 11ul. This PCR reaction was run under the same cycling temperatures described above.

Once we determined that the 5’UTR genomic and ORF7a subgenomic assays could be successfully duplexed, we then attempted to create a triplex assay by incorporating the N gene subgenomic assay into the duplex. This would allow us to efficiently quantitate two subgenomic RNA molecules alongside the genomic length RNA. Different primer concentrations of the common leader forward primer from 0.71, 1.43 and 2.86 µM were evaluated as part of the optimization of this assay. The final PCR reaction for the triplex assay was setup by adding 1x Brilliant Multiplex qPCR mastermix (Agilent), 2.86µM of the shared SARS-CoV2 leader forward primer, 0.71 µM of each of the 5’UTR and ORF7a and N-gene reverse primers, 0.14 µM of each of the 5’UTR, ORF7a and N-gene probes, 2ul of sample/control cDNA and nuclease free water to a final volume of 14ul. The triplex assay was run in duplicate under the same temperature conditions as described above. The PCR efficiency of the triplex reactions was determined by testing the serial dilution of the PCR amplicons as described above.

### Real time assays to quantitate positive and negative sense SARS-CoV-2 RNA

During replication of SARS-CoV-2 in cells, the virus produces negative sense RNA to act as a template for the production of full length copies of the virus genome length RNA and the various subgenomic RNAs. We had previously detected negative strand RNA in three of eight diagnostic swab samples^[23]^, however the quantity of negative strand SARS-CoV-2 RNA was 150 and 20 fold lower than positive strand genomic and subgenomic RNA respectively^[23]^, and therefore we could only detect it in samples with a very high virus load (low Ct). To create potentially more sensitive assays to detect strand specific RNA in more SARS-CoV-2 samples, we attempted to adapt the probe-based PCRs described above to single step, strand-specific PCR reactions utilizing sense specific primer reverse transcription followed by PCR.

We initially used the SARS-CoV-2 48 hr cell culture, for which we had ample RNA available, for the development and optimization of the strand based assays. For each strand specific PCR, 2µl of the RNA from a sample was denatured at 95°C for 3 mins and then rapidly cooled on ice. The denatured RNA was then mixed with 1x Brilliant II qRT-PCR 1-Step QRT-Master Mix (Agilent), 1µl of 10µM primer (the PCR forward primer for negative sense cDNA synthesis and the PCR reverse primer for positive sense cDNA synthesis), 0.5ul of RT/RNase block enzyme mixture (Agilent) and nuclease free water to a final volume of 8.8ul. This was then incubated at 50°C for 30 min followed by 95°C for 8 min. At this point, 1µl of 10uM complementary primer and 0.2µl of the 10µM stock of the corresponding probe were added to the reaction to bring the reaction volume to 10µl. This was then incubated at 95°C for 2 min, then 40 cycles of 95°C for 3 sec, 58°C for 30 sec and 64°C for 30 sec in a Quantstudio 6 real-time PCR thermocycler. Serial dilutions of the gel purified amplicons were used to determine the efficiency slope of each PCR reaction. During testing with the positive and negative controls, it was determined that the reverse primer of the 5’UTR PCR, while a relatively efficient PCR primer, was inefficient at synthesising first strand cDNA. Attempts to improve the cDNA synthesis, including trialling different reaction temperatures, adding primer at the denaturation step, using different RT enzymes or slightly different reverse primers for the cDNA step, were not successful in improving the performance of the positive sense-specific 5’-UTR PCR. The positive sense 5’UTR assay was therefore unreliable/not sensitive enough to quantitate the amount of positive sense genomic length RNA present within the samples. We therefore selected and used the sense-specific PCRs for the 7a total instead of the 5’UTR together with the 7a subgenomic and N subgenomic as three separate strand specific assays. These three assays performed as expected with efficiencies of around 95% similar to the non-strand specific versions of these assays.

Once we were confident that we had developed an efficient set of strand specific assays, we focussed on the samples which likely had enough virus RNA load to allow detection of strand specific RNA targets. Fourteen SARS-CoV-2 positive swabs, the 48 hour SARS-CoV-2 cell culture supernatant, and the negative individual and pooled negative swab sample 1 were tested in the strand specific assays described above. Some of the clinical samples had little RNA remaining at this point in the investigation, so all RNAs were diluted 1:4 in low TE prior to cDNA synthesis except for the cell culture, which was not diluted and the sample from individual 3 (GC-13), which was diluted 1:6. Due to the poor performance of the 5’ UTR positive sense PCR, ratios of the amount of strand specific subgenomic RNAs were estimated by the delta-Ct between the 7a/N subgenomic assays and the 7a-total assay, which was used as a proxy for the amount of genomic RNA present in the sample.

### Development of Ampliseq Mini-Panels to specifically detect SARS-CoV-2 subgenomic RNAs

We previously reported on the ability of the commercially available SARS-CoV-2 Ampliseq Panel from Thermofisher Scientific [https://www.thermofisher.com/au/en/home/life-science/sequencing/dna-sequencing/microbial-sequencing/microbial-identification-ion-torrent-next-generation-sequencing/viral-typing/coronavirus-research.html] to detect subgenomic RNAs ^[23]^ as part of SARS-CoV-2 genomic sequencing. This occurred as a result of the two forward primers within the leader sequence (the first two primers of each of the two commercial Ampliseq primer pools) of the SARS-CoV-2 genome producing subgenomic RNA specific amplicons with the reverse primers downstream of the Transcription Regulatory Sites (TRS) sequence in each of the different SARS-CoV-2 genes^[23]^. The method was sensitive and detected subgenomic RNAs in all samples tested despite these amplicons competing with the 242 SARS-CoV-2 and host gene specific amplicons within the Ampliseq reaction. To determine if the sensitivity, and possibly also the ability to compare levels of individual amplicons/subgenomic RNAs, of this method could be improved, we designed/selected two new Ampliseq mini-panels containing only the primers required to amplify the subgenomic RNA molecules, i.e. a forward sense primer sitting within the leader sequence and a reverse primer from each of the potential 10 SARS-CoV-2 subgenomic RNAs downstream of the predicted TRS sequences. A single reverse primer within the 5’UTR and a primer pair targeting the host cellular TATA box binding protein (TBP) mRNA were also included in each of the two Ampliseq panels to hopefully provide an estimate of the amount of SARS-CoV-2 genomic RNA and host cellular mRNA within the sample, respectively. The full list of primers in each Ampliseq mini-panel can be found in Supplementary Table S4 and were obtained from Thermofisher. Mini-panel 1 included the first primer from the commercial SARS-CoV-2 Ampliseq primer pool 1, which terminated at nucleotide 42 of SARS-CoV-2 Wuhan-Hu-1 (NC_045512.2). Mini-panel 2 included the first primer from the second SARS-CoV-2 Ampliseq primer pool which ended at nucleotide position 52 of NC_045512.2. The Ampliseq primers for these mini-panels were selected by us from the full SARS-CoV-2 Ampliseq Panel (Thermofisher Scientific) and obtained from the Ampliseq custom service team at Thermofisher Scientific.

Ampliseq reactions were performed using the random hexamer VILO cDNA prepared above, and the Ion Ampliseq Plus Library Kit (Thermofisher Scientific, Victoria Australia) as per manufacturer’s instructions initially with three modifications. The first was the addition of 2µM of SYTO 9 (Thermofisher Scientific, Victoria, Australia) to each reaction to enable the PCR amplification to be monitored in real time. The second was to initially increase the PCR cycles from 21 to 31 cycles which corresponded with most samples entering the exponential phase of the PCR amplification. The third was to decrease the individual primer concentration to 71nM (Total primer concentration 1µM). In a separate set of reactions, we used selected samples made into cDNA by either random hexamers (SuperScript™ VILO™ Master Mix, Thermofisher Scientific) or cDNA made with the Ampliseq mini-panel 1 primers using superscript IV Reverse transcriptase (Thermofisher Scientific) and 21 Ampliseq PCR reaction cycles. Sample libraries were quantitated using the Ion Library Taqman quantitation kit, pooled and loaded onto Ion Torrent 530 chips using an Ion Chef templating robot and sequenced using an Ion Torrent S5XL. Seven samples were also sequenced with the full SARS-CoV-2 Ampliseq Panel as per the manufacturer’s instructions using 21 cycles of PCR amplification and the reads assembled into SARS-CoV-2 consensus sequences using Ion Torrent plugins as described previously^[23]^ and analysed for the presence of subgenomic reads described briefly below. Generated sequences were then mapped to a custom SARS-CoV-2 subgenomic RNA reference which contained the sequence of the ORF1ab and the start of each subgenomic RNA molecule including the 65/69 nucleotides of the leader sequence, the TRS and the 5’ end of each of the genes of the SARS-CoV2 virus including up to where each reverse primer was predicted to anneal within that gene’s sequence^[23]^. Mapping of the reads was performed by TMAP software included in the Ion torrent Server software suite 5.10.1^[39]^. Counts of reads mapping to each subgenomic, genomic and host target were obtained by using the Coverage Analysis plugin (v5.12.0.0) on the Ion Torrent Server, using a minimum mapping quality score of 20 and minimum alignment length of 20. Read mapping counts were checked by visualizing the reads and coverage using Integrative Genome Viewer (IGV) 2.6.3^[40]^.

### Measuring the nucleic acid quantity and quality in the nasopharyngeal swab samples

To investigate the host RNA and host inflammatory response detectable in the nasopharyngeal mucosa to SARS-CoV-2 infection, as represented by the swab samples included in our study, we first measured the quantity and quality of the extracted nucleic acid using Agilent RNA 6000 Pico Chips on an Agilent 2100 bioanalyzer (Agilent Technologies, California, USA) which calculated the RNA integrity number (RIN) from the electropherogram of each sample. We then performed a Kendall Rank correlation analysis between the total nucleic acid quantity (pg/µl) and both the RIN and the Ct’s of the non-strand specific single target and triplex PCR assays, and the strand specific assays.

## Data Availability

The sequence reads of our SARS-CoV-2 positive and negative samples reported here have been deposited along with their corresponding fasta reference files in the NCBI Sequence Read Archive (SRA) under BioProjectID: PRJNA738539. Assembled near full length SARS-CoV-2 genomic sequences are deposited in NCBI GenBank under accessions MZ410617-MZ410622.
Other assembled nucleotide sequences or nucleotide sequence read archives mentioned are publicly available at NCBI ([https://www.ncbi.nlm.nih.gov/nuccore/ and https://www.ncbi.nlm.nih.gov/sra/]. All other data supporting the findings of this manuscript are available in the Supplementary Information file or from the corresponding author upon reasonable request. Source data are provided with this paper.

## Acknowledgements

We gratefully acknowledge clinical staff for sample collection and the diagnostic staff at the Australian Rickettsial Reference Laboratory (ARRL) and Australian Clinical Labs (ACL) for performing the initial diagnostic SARS-CoV-2 testing, in particular Professor John Stenos and Dr Mythili Tadepalli from ARRL and Professor Owen Harris, Dr Kwee Chin Liew and Dr Richard McCoy from ACL for their assistance in testing, aliquoting, storing and providing samples. We acknowledge Dr Darcie Paige Cooper for retrieving summary clinical information and Professors Peter Vuillermin and Eugene Athan for the coordination of the Barwon Health and Deakin University COVID-19 Research Task Force and Cohort Study and the human research ethics approval. We acknowledge Thermofisher Scientific, Victoria, Australia, for supplying the SARS-CoV-2 sequencing panel and for help with the custom Ampliseq mini-panels used. We acknowledge colleagues at the CSIRO Australian Centre for Disease Preparedness, especially Dr Alexander J. McAuley and Professor Seshadri S. Vasan, for providing the SARS-CoV-2 positive cell culture. We acknowledge Professor Alister Ward and Dr Poshmaal Dhar for their helpful comments on the draft manuscript. We also acknowledge Dr Jason Hodge, laboratory manager of the GCEID laboratory for his technical input. This research was funded by Deakin University, Barwon Health and CSIRO and from the National Health and Medical Research Council (NHMRC) equipment grant number GNT9000413 to S.A.

## Author Contributions

S.A. initiated the work and coordinated work carried out at the GCEID laboratory. T.R.B. and S.A. designed the PCRs and primers/probes and S.A. selected and designed the mini-panels for NGS. T.R.B. managed the collection of samples and performed all strand and non-strand specific PCR assays. A.C. performed all mini-panel, SARS-CoV-2, bioanalyzer and statistical analyses. S.A., A.C. and T.R.B. all analysed the PCR and sequence data. A.C. prepared the initial draft of the paper together with S.A. and all authors contributed to the editing and drafting process.

## Additional Information

### Data Availability

The sequence reads of our SARS-CoV-2 positive and negative samples reported here have been deposited along with their corresponding fasta reference files in the NCBI Sequence Read Archive (SRA) under BioProjectID: PRJNA738539. Assembled near full length SARS-CoV-2 genomic sequences are deposited in NCBI GenBank under accessions MZ410617-MZ410622. Other assembled nucleotide sequences or nucleotide sequence read archives mentioned are publicly available at NCBI ([https://www.ncbi.nlm.nih.gov/nuccore/ and https://www.ncbi.nlm.nih.gov/sra/]. All other data supporting the findings of this manuscript are available in the Supplementary Information file or from the corresponding author upon reasonable request. Source data are provided with this paper.

### Code Availability

Not applicable.

## Supplementary Materials

The following is available online; Supplementary Information file 1 including Supplementary Tables S1-4 and Supplementary Figures S1-8. Supplementary data is supplied as a separate Excel file.

