## Supplementary Information for "Subgenomic and negative sense RNAs are not markers of active replication of SARS-CoV-2 in nasopharyngeal swabs"

Supplementary Table S1. Ct values obtained from the single target SARS-CoV-2 genomic and subgenomic PCR assays for the swab samples from the 16 SARS-CoV-2 positive individuals, negative individual and pool and from 48hr virus cell culture supernatant.

| **Individual** | **Swab Sample ID** | **Days since onset of symptoms/Initial swab** | **5'UTR Genomic PCR** | **ORF7a subgenomic PCR** | **N gene subgenomic PCR** | | **7a Genomic and Subgenomic PCR** |
|---|---|---|---|---|---|---|---|
| 1 | GC-28 | 0* (SARS-CoV-2 NEG) | ND | ND | NEG | NEG | ND |
| 2 | GC-26 | 7 | 25.6 | 30 | 27.6 | 27.4 | 25.3 |
| 3 | GC-13 | 4 | 29.6 | 33.3 | 29.8 | 30.1 | 28.6 |
| 4 | GC-24 | 14* | 35.4 | NEG | 36.2 | NEG | 33.6 |
| 6 | GC-14 | 0 | 18.7 | 23.7 | 20.2 | 19.8 | 17.9 |
| 6 | GC-23 | 11 | 33.7 | 38.6 | 34.7 | 34.6 | 32.6 |
| 6 | GC-51 | 17 | 39.8 | NEG | 34.1 | 35.5 | 36 |
| 7 | GC-21 | 16 | 35.7 | NEG | 36.7 | NEG | 33.7 |
| 7 | GC-58 | 47 | NEG | NEG | NEG | NEG | NEG |
| 8 | GC-25 | 2 | 21.5 | 25.4 | 21.1 | 20.9 | 19.9 |
| 11 | GC-242 | 0 | 27.3 | 30.3 | 26.5 | 26.7 | 26.1 |
| 11 | GC-290 | 11 | 33.5 | 36.7 | 33.5 | 34.2 | 31.9 |
| 12 | GC-251 | 0 | 17 | 21.2 | 17.6 | 17.6 | 16.5 |
| 12 | GC-288 | 10 | 36.7 | 37.1 | 34.8 | NEG | 34.7 |
| 13 | GC-277 | 0 | 20.2 | 25.1 | 21.4 | 21.2 | 18.9 |
| 13 | GC-329 | 19 | 37 | NEG | 35.7 | NEG | 34.6 |
| 14 | GC-238 | 0 | 24.3 | 29.5 | 26.7 | 26.7 | 22.7 |
| 14 | GC-291 | 13 | 22 | 24.8 | 22.8 | 23 | 20.4 |
| 16 | GC-366 | 14 | 39 | NEG | 31.9 | 38.5 | 33.7 |
| 16 | GC-365 | 17 | 38.1 | NEG | NEG | NEG | 34.1 |
| 17 | GC-316 | 1 | 19.5 | 22.6 | 19.5 | 19.6 | 18.1 |
| 18 | GC-310 | 17 | 21.5 | 25.9 | 23.7 | 23.8 | 21.7 |
| 19 | GC-199 | 1 | 23 | 28.1 | 24.8 | 24.8 | 23.1 |
| 20 | GC-292 | 10 | 17.6 | 20.3 | 17.5 | 17.3 | 17.2 |
| 21 | GC-243 | 0 | 24.7 | 27.6 | 23.4 | 23.7 | 23.4 |
| SARS-CoV-2 Negative swab pool | Pool of 10 Neg. swabs | Not specified | ND | ND | NEG | NEG | ND |
| Cell culture 48Hr | | 48hrs post inoculation | 16.8 | 20.4 | 19.5 | 19.3 | ND |

ND: Test not performed. NEG: SARS-CoV-2 not detected.

*Date of onset of clinical signs not provided. Number of days since the collection of the first nasopharyngeal swabs is shown.

†Sample exhausted

Supplementary Table S2. Ct values obtained from the duplex SARS-CoV-2 genomic and subgenomic PCR assays for the swab samples from the 16 SARS-CoV-2 positive individuals, negative individual and pool and from 48hr virus cell culture supernatant.

| **Individual** | **Swab Sample ID** | **Days since onset of symptoms/Initial swab** | **5'UTR Genomic Duplex PCR** | **ORF7a subgenomic Duplex PCR** |
|---|---|---|---|---|
| 1 | GC-28 | 0* | NEG | NEG |
| 2 | GC-26 | 7 | 23.9 | 27.5 |
| 3 | GC-13 | 4 | 27.3 | 30.7 |
| 4 | GC-24 | 14* | 33.8 | NEG |
| 6 | GC-14 | 0 | 16.8 | 20.5 |
| 6 | GC-23 | 11 | 31.6 | NEG |
| 6 | GC-51 | 17 | 35.4 | NEG |
| 7 | GC-21 | 16 | 34.5 | NEG |
| 7 | GC-58 | 47 | NEG | NEG |
| 8 | GC-25 | 2 | 19 | 22.4 |
| 11 | GC-242 | 0 | 25.8 | 28.6 |
| 11 | GC-290 | 11 | 31.9 | 33.9 |
| 12 | GC-251 | 0 | 15.7 | 18.9 |
| 12 | GC-288 | 10 | 33.7 | NEG |
| 13 | GC-277 | 0 | 18.3 | 22.4 |
| 13 | GC-329 | 19 | 34.8 | 36.6 |
| 14 | GC-238 | 0 | 21.8 | 27.6 |
| 14 | GC-291 | 13 | 19.9 | 23.3 |
| 16 | GC-366 | 14 | 34.7 | NEG |
| 16 | GC-365 | 17 | 34.7 | NEG |
| 17 | GC-316 | 1 | 17 | 20.3 |
| 18 | GC-310 | 17 | 20.5 | 24.4 |
| 19 | GC-199 | 1 | 20.6 | 25.6 |
| 20 | GC-292 | 10 | 15.4 | 18.9 |
| 21 | GC-243 | 0 | 22.7 | 25.6 |
| SARS-CoV-2 Negative swab pool | Pool of 10 Neg. swabs | Not specified | ND | ND |
| Cell culture 48Hr | | 48 hrs post inoculation | 17.3 | 20 |

ND: Test not performed. NEG: SARS-CoV-2 not detected.

*Date of onset of clinical signs not provided. Number of days since the collection of the first nasopharyngeal swabs is shown.

Supplementary Table S3. Table showing the sequence and position of each of the primers and probes of the SARS-CoV-2 genomic and subgenomic PCR assays.

| Set | Primers/ Probes | Sequence (5′->3′) | Length | Region | Amplification |
|---|---|---|---|---|---|
| 1 | SARS-CoV-2-Leader-SA-45-66-F | GATCTCTTGTAGATCTGTTCTC | 22 | Leader | 5'UTR genomic |
| | SARS-CoV-2_TRB_RP4_206_187 | GACGAAACCGTAAGCAGCCT | 20 | 5′ UTR | |
| | SARS-CoV-2_TRB_ 5'UTR Sense_148-172 (Cy5) | TGTCGTTGACAGGACACGAGTAACT | 25 | 5′ UTR | |
| 2 | SARS-CoV-2-Leader-SA-45-66-F | GATCTCTTGTAGATCTGTTCTC | 22 | Leader | ORF-7a sub-genomic |
| | SARS-CoV-2_TRB_RP1-3-27531-27512 | AAATGGTGAATTGCCCTCGT | 20 | Orf7a | |
| | SARS-CoV-2_TRB_ ORF7a Sense_27428-27455 (VIC) | TCGCTACTTGTGAGCTTTATCACTACCA | 28 | Orf7a | |
| 3 | SARS-CoV-2_TRB_FP2-27401-27425 | TTATTCTTTTCTTGGCACTGATAAC | 25 | Orf7a | ORF-7a total |
| | SARS-CoV-2_TRB_RP1-3-27531-27512 | AAATGGTGAATTGCCCTCGT | 20 | Orf7a | |
| | SARS-CoV-2_TRB_ ORF7a Sense_27428-27455 (VIC) | TCGCTACTTGTGAGCTTTATCACTACCA | 28 | Orf7a | |
| 4 | SARS-CoV-2-Leader-SA-45-66-F | GATCTCTTGTAGATCTGTTCTC | 22 | Leader | N gene sub-genomic |
| | RP1-SCV2-N-28396-28378-TRB | CCGACGTTGTTTTGATCGC | 19 | N gene | |
| | SARS-CoV-2_TRB_N-Probe_Sense_28330-28355 (FAM) | ACCCTCAGATTCAACTGGCAGTAACC | 26 | N gene | |

Supplementary Table S4. The names of primers selected from the Ampliseq SARS-CoV-2 Panel amplicons to make the two mini-panels. The 3' end position of each primer is listed in relation to the sequence of SARS-CoV-2 Wuhan-Hu-1 (NC_045512.2). The primer sequence is proprietary of Thermofisher Scientific.

**Mini Panel 1 primers**

| Target | Primer name | Location on SARS-CoV-2 genome (NC_045512.2) |
|---|---|---|
| 5' UTR Leader (Forward) | R1_1.1363702 F | 42 |
| 5' UTR – genomic (Reverse) | R1_1.1363702 R | 229 |
| S gene (Reverse) | R1_1.22.714732 R | 21664 |
| ORF3a (Reverse) | R1_1.26.628976 R | 25477 |
| E gene (Reverse) | R1_1.27.887996 R | 26307 |
| M gene (Reverse) | R1_1.27.1073732 R | 26540 |
| ORF 6 (Reverse) | R1_1.28.162880 R | 27095 |
| ORF 7a (Reverse) | R1_1.28.1321347 R | 27453 |
| ORF 7b (Reverse) | R1_1.28.99230 R | 27701 |
| ORF 8 (Reverse) | R1_1.28.958844 R | 27918 |
| N gene (Reverse) | R1_1.29.157415 | 28336 |
| ORF10 (Reverse) | R2_1.79363 (Reverse complement) | 29593* |

**Mini panel 2 primers**

| Target | Primer name and location on SARS-CoV-2 genome | Location on SARS-CoV-2 genome (NC_045512.2) |
|---|---|---|
| 5' UTR Leader (Forward) | R1_1.1.80142 F | 52 |
| 5' UTR – genomic (Reverse) | R1_1.1363702 R | 229 |
| S gene (Reverse) | R1_1.22.714732 R | 21664 |
| ORF3a (Reverse) | R1_1.26.628976 R | 25477 |
| E gene (Reverse) | R1_1.27.887996 R | 26307 |
| M gene (Reverse) | R1_1.27.1073732 R | 26540 |
| ORF 6 (Reverse) | R1_1.28.162880 R | 27095 |
| ORF 7a (Reverse) | R1_1.28.1321347 R | 27453 |
| ORF 7b (Reverse) | R1_1.28.99230 R | 27701 |
| ORF 8 (Reverse) | R1_1.28.958844 R | 27918 |
| N gene (Reverse) | R1_1.29.157415 | 28336 |
| ORF10 (Reverse) | R2_1.79363 (Reverse complement) | 29593* |

*The ORF10 primer was the reverse complement of the original forward primer of this amplicon, and so its 5' end position is provided. We estimate that the primer is 30nt long

Supplementary Figure S1. The Ct values of the four individual PCR assays targeting the genomic 5' UTR, subgenomic ORF7a and N gene, and total ORF7a (detecting genomic and S, ORF3a, E, M, ORF6 and ORF7a subgenomic RNAs) for the naso-oropharyngeal swabs from SARS-CoV-2 positive individuals, the 48 hr cell culture and the SARS-CoV-2 negative pool of swabs. The results are ordered by ascending values of the 5'UTR Ct and show that both the ORF7a and N gene subgenomic, and the ORF7a total PCR Ct's also increase as the UTR Ct increases. Above a Ct of around 30 for the 5'UTR, the detection of subgenomic RNAs becomes less consistent/below levels of detection/quantitation. (n=25 data points from 25 biological samples run once (Subgenomic N run twice)).

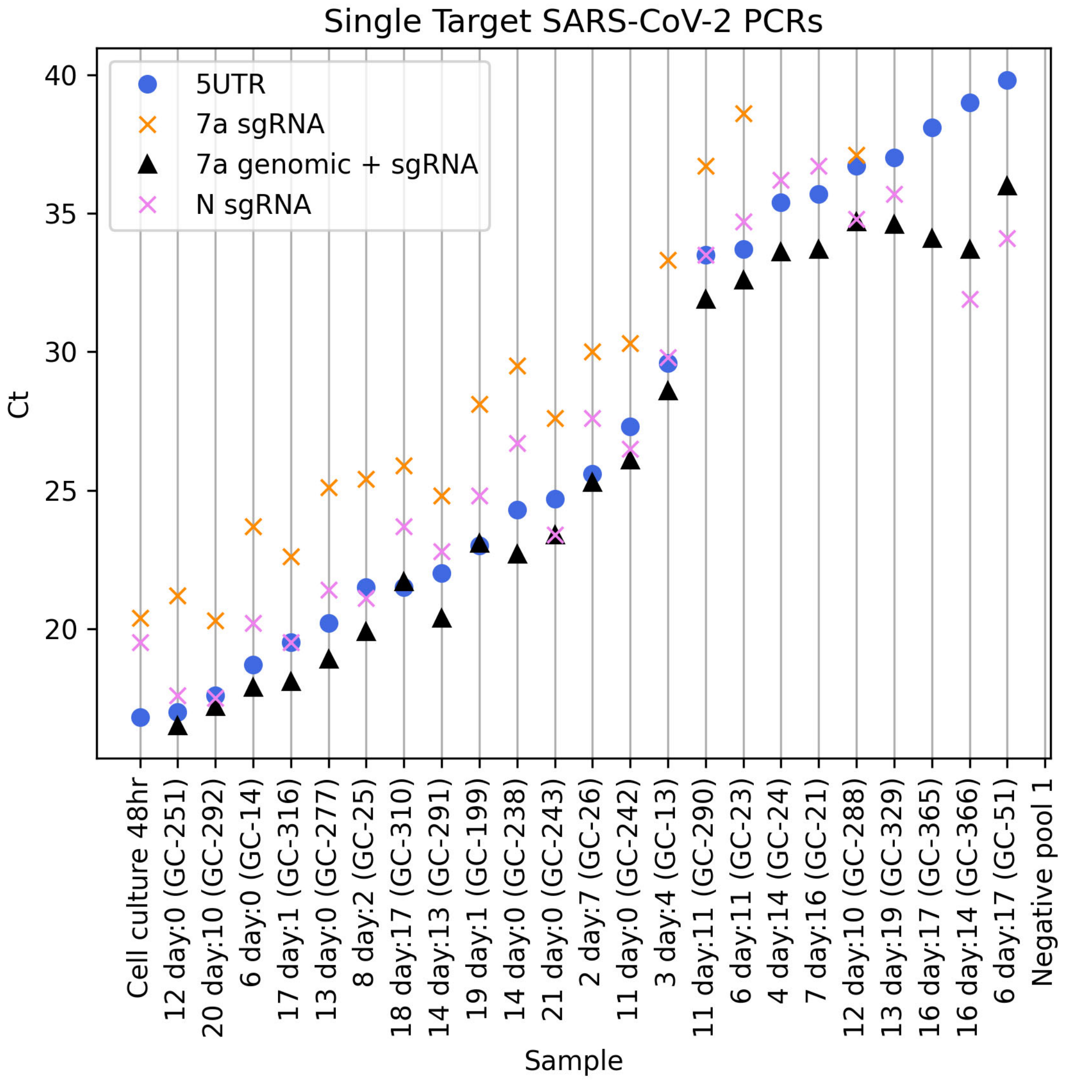

Supplementary Figure S2. Duplex PCR results showing the PCR Ct's for the genomic 5'UTR and the ORF7a subgenomic PCRs. The subgenomic ORF7a PCR was typically 2 to 4 Cts higher than the 5'UTR PCR. Above a 5'UTR Ct of 30, the subgenomic ORF7a assay was not able to consistently detect subgenomic targets. (n=25 data points from 25 biological samples run once)

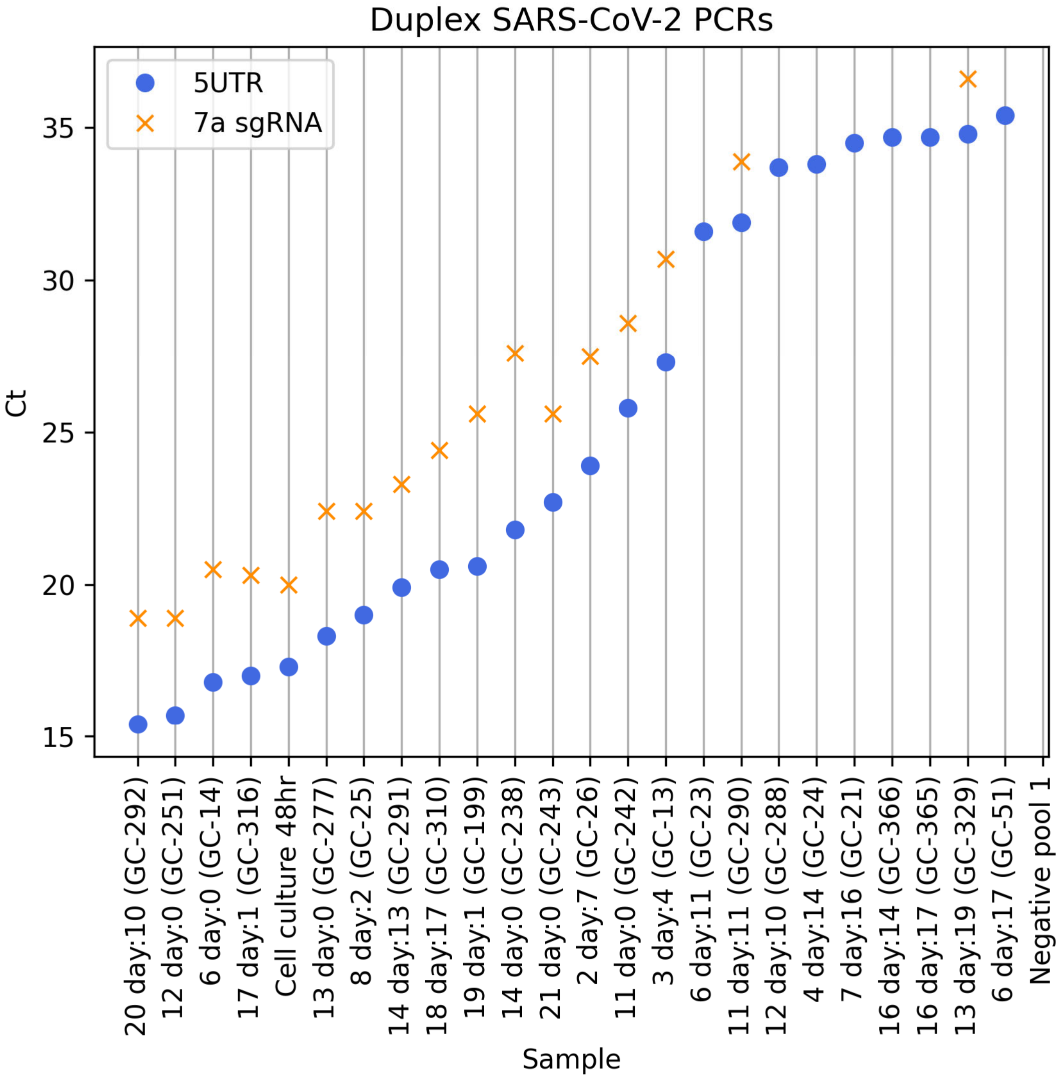

Supplementary Figure S3. Scatter plots of the strand specific RNA PCR Ct's and the time between symptom onset and swab collection

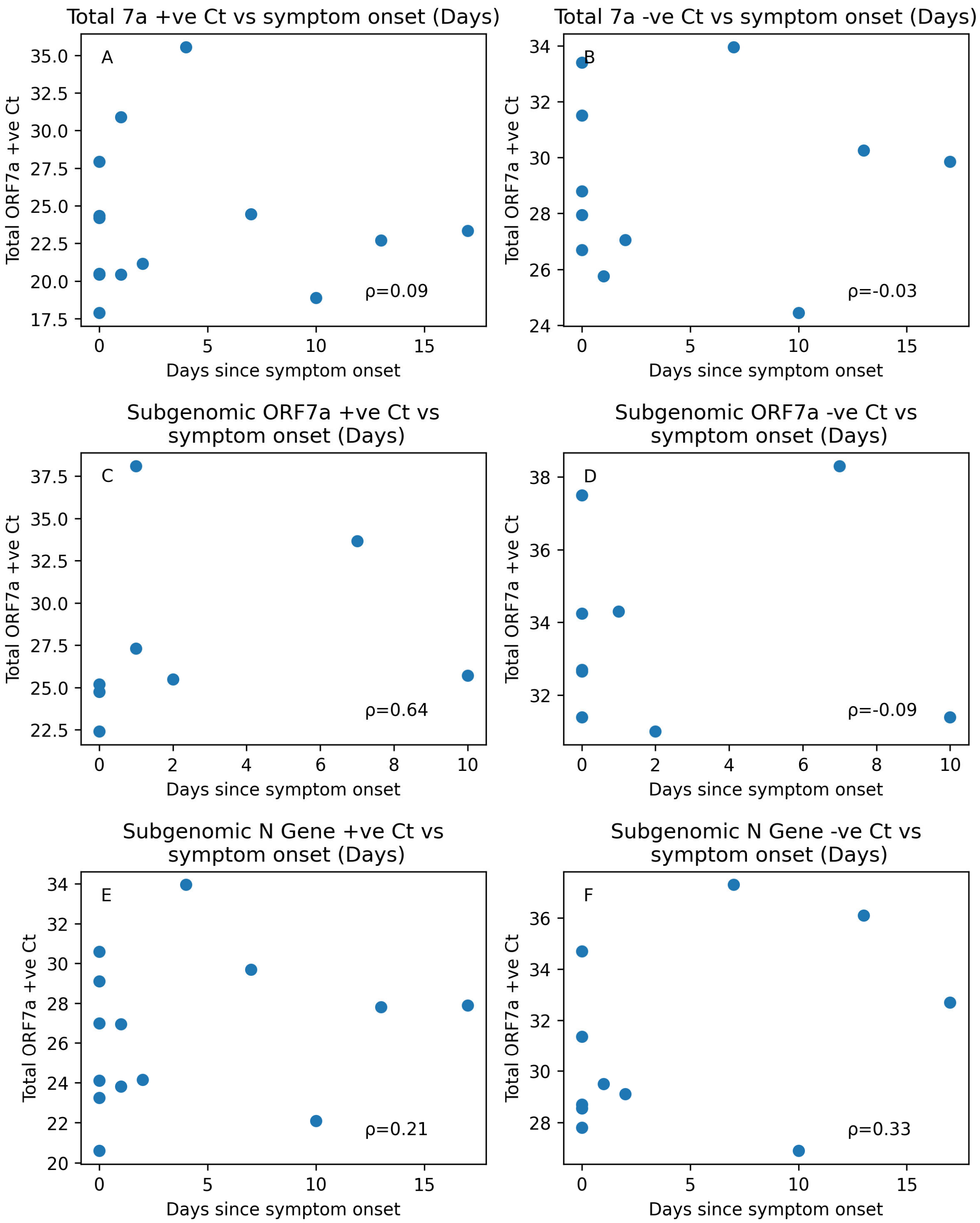

Supplementary Figure S4. Relative percentage of reads mapped to each subgenomic RNA using the Ampliseq mini-panel 1, cDNA synthesis with random hexamers and 21 PCR amplification cycles. (n=8 data points from 8 biological samples run once)

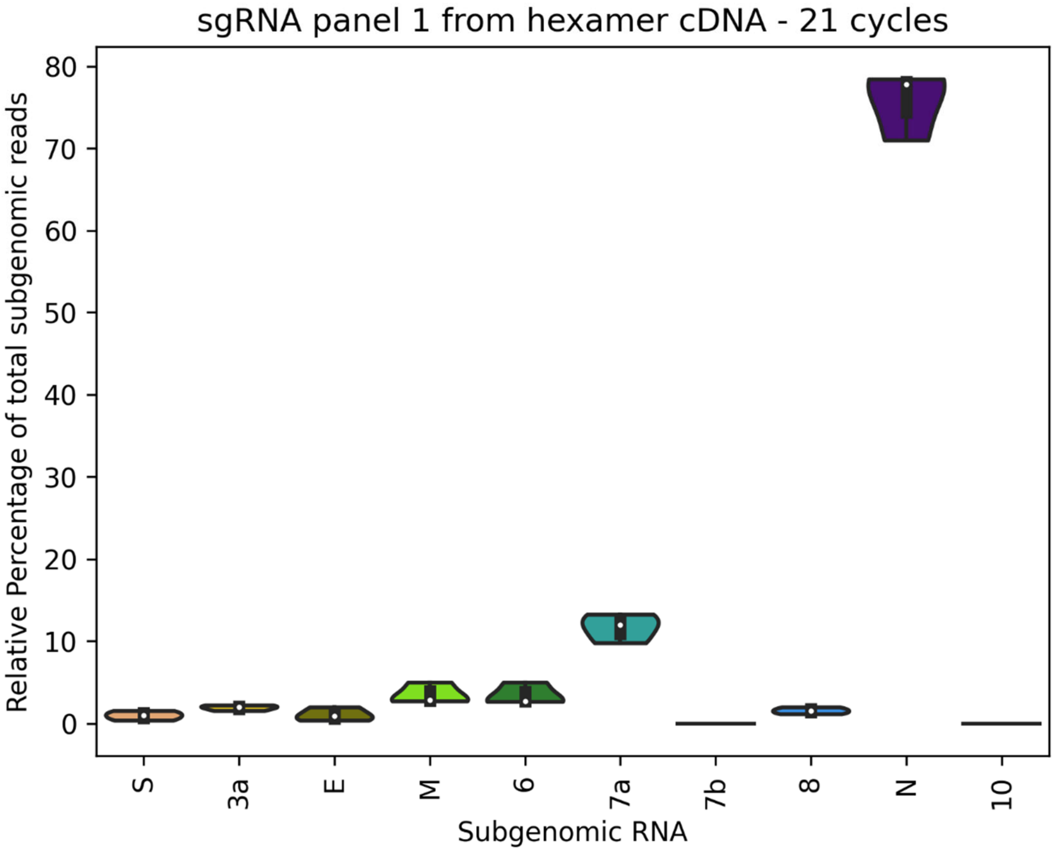

Supplementary Figure S5. Relative percentage of reads mapped to each subgenomic RNA using the Ampliseq mini-panel 2, cDNA synthesis with random hexamers and 31 PCR amplification cycles. (n=25 data points from 25 biological samples run once)

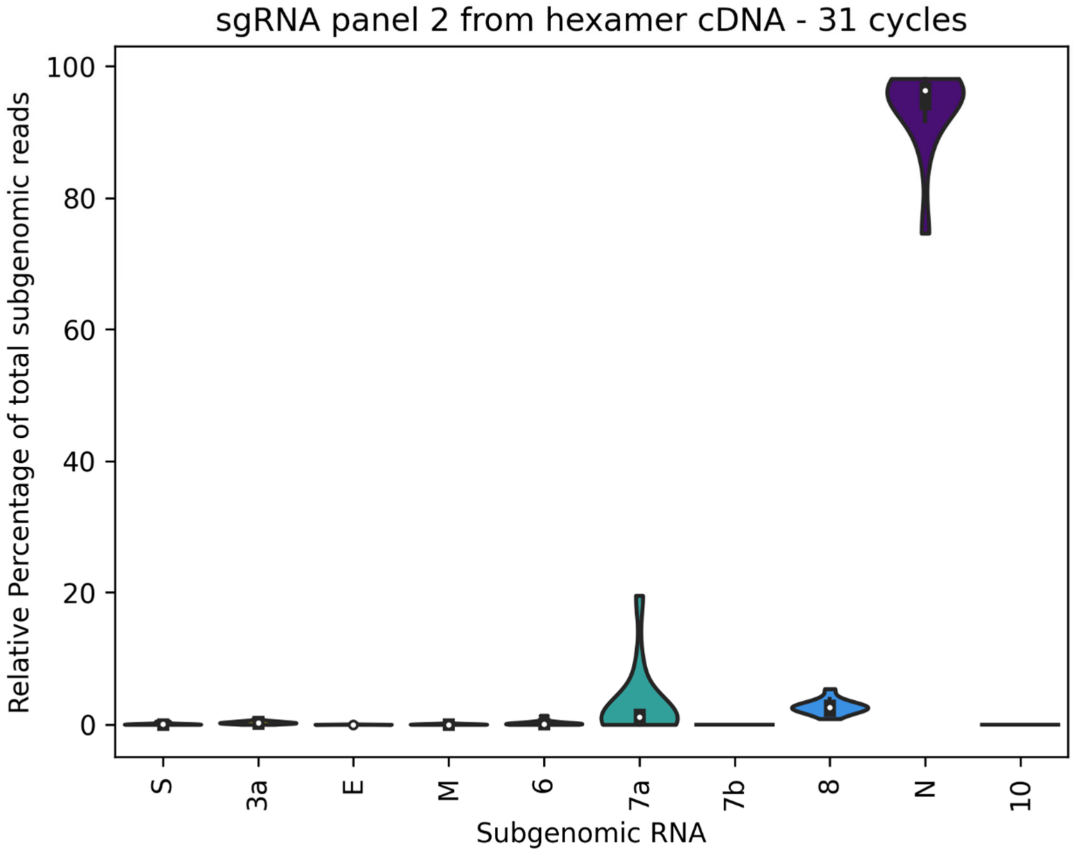

Supplementary Figure S6. The relative percentage of subgenomic reads from the full SARS-CoV-2 Ampliseq panel.(n=7 data points from 7 biological samples run once)

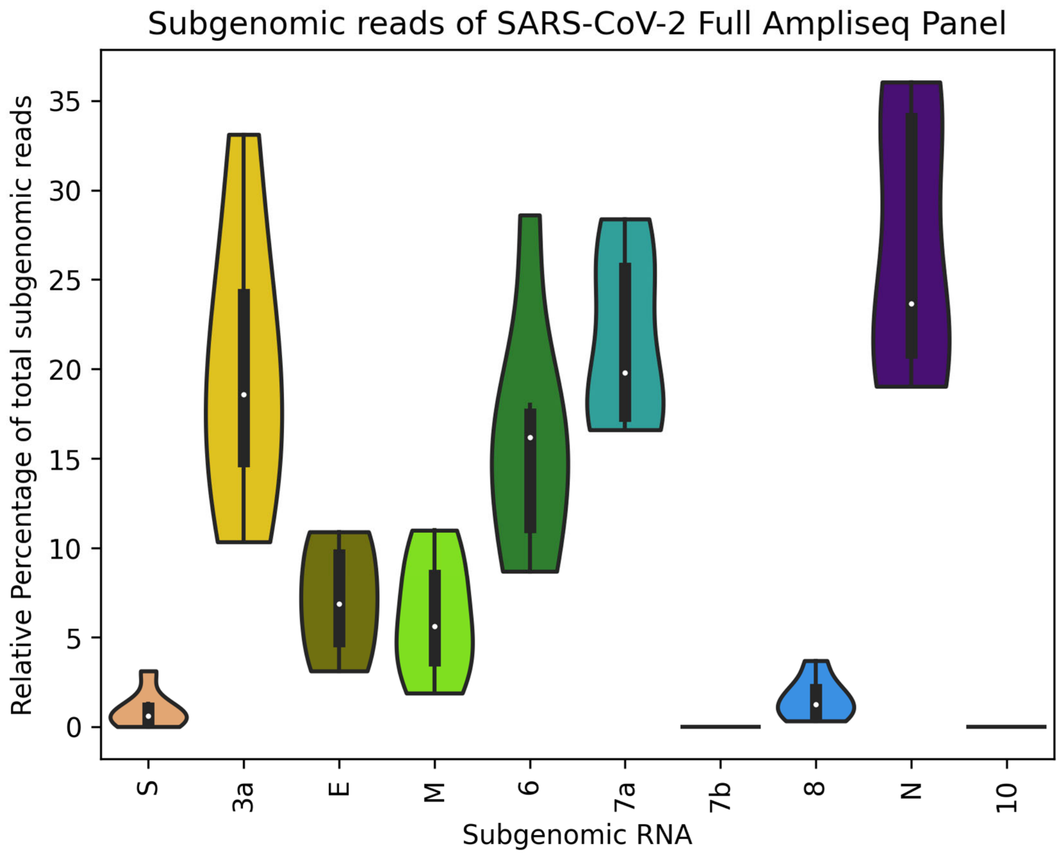

Supplementary Figure S7. Plot of the triplex 5'UTR PCR assay vs the quantity of RNA (pg/µl) present within a sample as measured by an RNA Pico 6000 chip on an Agilent Bioanalyzer 2100. (n=21 from 21 biological samples run once)

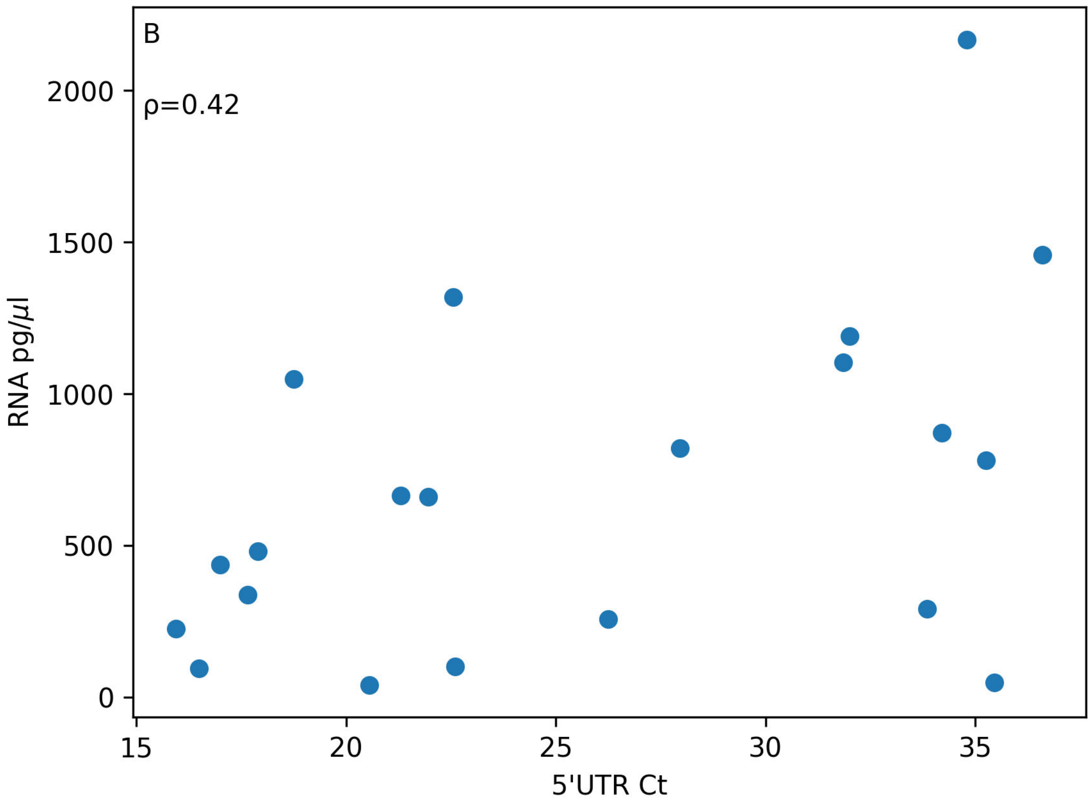

Supplementary Figure S8. The Ct of the triplex SARS-CoV-2 PCRs by the RNA integrity number (RIN) measured by the Bioanalyzer. Samples were divided into three groups, those with no RIN or a RIN<3, those with a RIN between 3 and 6, and those with a high RIN score (>6). Only one nasopharyngeal swab sample had a high RIN. The PCR Ct's showed no relationship with the RIN.

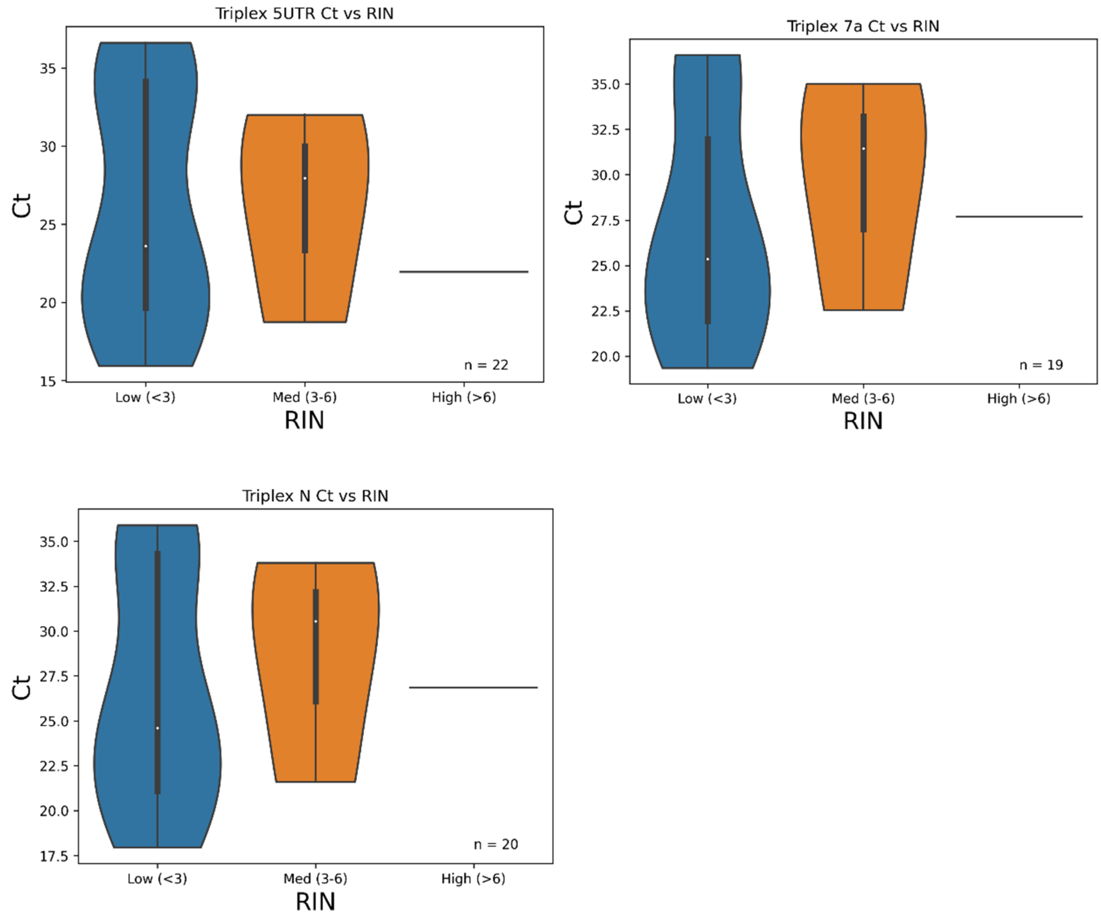
